# Area-level social and structural inequalities determine mortality related to COVID-19 diagnosis in Ontario, Canada: a population-based explanatory modeling study of 11.8 million people

**DOI:** 10.1101/2022.03.14.22272368

**Authors:** Linwei Wang, Andrew Calzavara, Stefan Baral, Janet Smylie, Adrienne K. Chan, Beate Sander, Peter C. Austin, Jeffrey C. Kwong, Sharmistha Mishra

## Abstract

**Importance:** Social determinants of health (SDOH) play an important role in COVID-19 outcomes. More research is needed to quantify this relationship and understand the underlying mechanisms.

**Objectives:** To examine differential patterns in COVID-19-related mortality by area-level SDOH accounting for confounders; and to compare these patterns to those for non-COVID-19 mortality, and COVID-19 case fatality (COVID-19-related death among those diagnosed).

**Design, setting, and participants:** Population-based retrospective cohort study including all community living individuals aged 20 years or older residing in Ontario, Canada, as of March 1, 2020 who were followed through to March 2, 2021.

**Exposure:** SDOH variables derived from the 2016 Canada Census at the dissemination area-level including: median household income; educational attainment; proportion of essential workers, racialized groups, recent immigrants, apartment buildings, and high-density housing; and average household size.

**Main outcomes and measures:** COVID-19-related death was defined as death within 30 days following, or 7 days prior to a positive SARS-CoV-2 test. Cause-specific hazard models were employed to examine the associations between SDOH and COVID-19-related mortality, treating non-COVID-19 mortality as a competing risk.

**Results:** Of 11,810,255 individuals included, 3,880 (0.03%) died related to COVID-19 and 88,107 (0.75%) died without a positive test. After accounting for demographics, baseline health, and other SDOH, the following SDOH were associated with increased hazard of COVID-19-related death (hazard ratios [95% confidence intervals]) comparing the most to least vulnerable group): lower income (1.30[1.09-1.54]), lower educational attainment (1.27[1.10-1.47]), higher proportion essential workers (1.28[1.10-1.50]), higher proportion racialized groups (1.42[1.16-1.73]), higher proportion apartment buildings (1.25[1.11-1.41]), and larger vs. medium household size (1.30[1.13-1.48]). In comparison, areas with higher proportion racialized groups were associated with a lower hazard of non-COVID-19 mortality (0.88[0.85-0.92]). With the exception of income, SDOH were not independently associated with COVID-19 case fatality.

**Conclusions and relevance:** Area-level social and structural inequalities determine COVID-19-related mortality after accounting for individual demographic and clinical factors. COVID-19 has reversed the pattern of lower non-COVID-19 mortality by racialized groups. Pandemic responses should include prioritized and community-tailored intervention strategies to address SDOH that mechanistically underpin disproportionate acquisition and transmission risks and shape barriers to the reach of, and access to prevention interventions.

**Key points:** *Question:* Are area-level social determinants of health factors independently associated with coronavirus disease 2019 (COVID-19)-related mortality after accounting for demographics and clinical factors?

*Findings:* In this population-based cohort study including 11.8 million adults residing in Ontario, Canada and 3,880 COVID-19-related death occurred between Mar 1, 2020 and Mar 2, 2021, we found that areas characterized by lower SES (including lower income, lower educational attainment, and higher proportion essential workers), greater ethnic diversity, more apartment buildings, and larger vs. medium household size were associated with increased hazard of COVID-19-related mortality compared to their counterparts, even after accounting for individual-level demographics, baseline health, and other area-level SDOH.

*Meaning:* Pandemic responses should include prioritized and community-tailored intervention strategies to address SDOH that mechanistically underpin inequalities in acquisition and transmission risks, and in the reach of, and access to prevention interventions.

## INTRODUCTION

Increasing evidence has confirmed the central role of social determinants of health (SDOH) in shaping variations in COVID-19 disease burden and severity(1, 2). Across high income settings, rates of COVID-19 diagnoses and deaths have been consistently correlated with socioeconomic status (SES)(3), and disproportionately affecting racialized groups(4-6).

In the context of infectious disease transmission, social and structural inequalities may shape differential health outcomes through the following pathway: differential susceptibility (e.g., modulated by the correlation between age and SDOH), contact patterns and networks (e.g., associated with occupation and housing conditions)(7, 8), reach/uptake of prevention interventions (e.g., differential access to testing(8, 9), effective isolation and quarantine(10), ability to reduce non-household contacts in the context of restrictions such as stay-at-home orders (11), access to vaccines(12)); and quality of treatment of illness (e.g., differential access to care).

To date, most studies have focused on SDOH such as SES as a composite index(9, 13, 14) and proxies for structural racism (race/ethnicity)(4, 6, 15). Few studies have examined other SDOH such as educational attainment, occupation and housing status; and even fewer have examined several SDOH in conjunction(1, 2). Moreover, studies on the relationship between SDOH and COVID-19 death were often conducted among diagnosed cases, or hospitalized populations(3). Although outcomes such as case fatality among diagnosed cases, and mortality while hospitalized provided important information regarding disease severity by SDOH, they were prone to selection biases, as SDOH were often correlated with exposure/infection, testing, diagnosis, and hospitalization(3, 13, 15).

In Canada, provisional Vital Statistics – Deaths data suggest that among COVID-19-related deaths occurred between January and August 2020, there were differences of 15 to 30 deaths per 100,000 in age-standardized COVID-19-related mortality, comparing residents of large urban vs. outside urban centers, lowest vs. highest income areas, highest vs. lowest ethno-cultural concentration, and residents of apartment buildings vs. detached homes(16). However, existing studies were not able to account for potential confounders such as baseline health conditions. Moreover, to date, no studies have estimated COVID-19-related mortality for SDOH subgroups while at the same time accounting for mortality unrelated to COVID-19, which is a competing risk for COVID-19-related mortality(17). Such an inquiry also provide opportunity to understand whether or not the same patterns of inequities drive both COVID-19 and non-COVID-19 related mortality.

Using population-based data among 11.8 million adults in Ontario, Canada’s most populous province, we examined differential patterns in COVID-19-related mortality across a set of area-level SDOH including SES (income, educational attainment, and occupation), ethnic diversity (proportion racialized groups, recent immigrations) and housing conditions (proportion apartment buildings, high-density housing, household size). We assessed whether differential patterns in COVID-19-related mortality by SDOH can be explained by demographics, baseline health (comorbidities and prior health care use), and other SDOH. We also compared patterns by SDOH in COVID-19-related mortality versus those in non-COVID-19 mortality, and in COVID-19 case fatality.

## METHODS

### Study design and subjects

We conducted a population-based retrospective cohort study of all community-dwelling adults in Ontario, Canada, a setting with universal health care (18). All individuals aged 20 years or older residing in Ontario as of March 1, 2020 and having a valid health card were identified using the Registered Persons Database (RPDB), and followed through March 2, 2021. We excluded residents in long-term care homes because they are not well represented in Canadian census data from which SDOH variables were determined(19, 20). We followed the Strengthening the Reporting of Observational Studies in Epidemiology (STROBE) reporting guideline. Data use in this project was authorized under Section 45 of Ontario’s Personal Health Information Protection Act, which does not require review by a Research Ethics Board.

### Outcomes

Our primary outcome of interest was COVID-19-related death, defined as death within 30 days following, or 7 days prior to a positive SARS-CoV-2 test. SARS-CoV-2 test result and date were determined based on records in Ontario Laboratory databases, and the COVID-19 surveillance system (Case and Contact Management System (CCM)). Death and date of death were determined using CCM and the RPDB. We estimated that use of both CCM and RPDB capture 99.3% of COVID-19-related death (**Appendix-Table-1**). Secondary outcome was non-COVID-19 death, defined as death without any history of a positive SARS-CoV-2 test.

We restricted our analyses to COVID-19-related deaths observed up to March 2, 2021 and cases diagnosed prior to Jan 31, 2021. As such, our analyses capture the first and second waves of regional pandemic with the majority (>95%) of cases representing the main strain of virus or alpha variant (21, 22).

### Covariates

Based on available data and existing literature(3, 4, 6, 8, 23, 24), we developed a conceptual framework to select SDOH variables, and potential confounders for the relationship between SDOH and outcomes, as hypothesized along the risk pathway of COVID-19-related mortality, including risk of infection, risk of testing if infected, and risk of death if diagnosed; with rationales of variable selection detailed in **Figure-1**.

**Figure 1.**
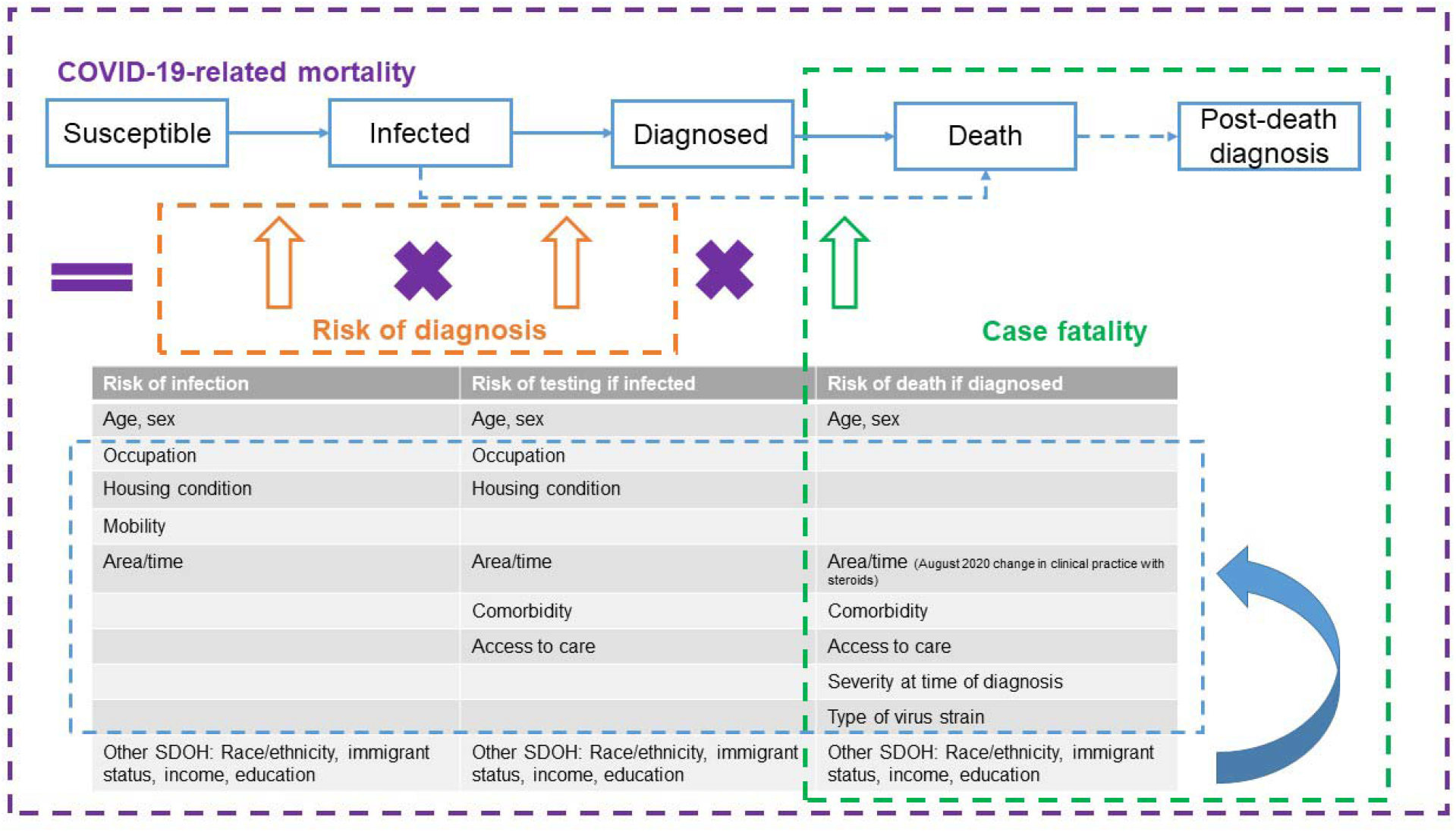
Conceptualization of risk factors for COVID-19-related mortality. Occupation (e.g., essential workers) might reflect contact rates at work; housing conditions (e.g.., living in apartment buildings vs detached houses, living in high-density housing, household size) and areas (e.g., urban vs. rural; public health unit) where an individual resides might reflect contact rates in communities; and therefore be associated with risk of infection and testing(1, 2). Individual’s baseline health (e.g., comorbidities and prior healthcare use) have been correlated with severity of infection and therefore associated with probability of testing and death(3). Socioeoconomic status, including household income and education, might affect exposure to the virus through working or living conditions, while also reflecting access to healthcare services, and therefore be associated with risk of infection, testing and death(4). Ethnic diversity, including racialized groups and recent immigration might be subject to systemic racism and socioeconomic inequalities, and affecting the risk pathway of COVID-related mortality(5, 6) Based on the conceptualized factors, we sourced data where available, at individual-level, otherwise at area-level. We don’t have data on mobility, however, we assume mobility may be a mediator between the relationship between SDOH and risk of infection. We don’t have clinical data on severity at time of diagnosis; however, we assume factors associated with risk of testing if infected will be associated with severity at time of diagnosis, and therefore, these factors were included in the models of case fatality. We don’t have data on type of virus strain; however, we restricted our analysis to first and second waves of regional pandemic before variants of concern took hold. SDOH: social determinants of health.

Our primary covariates of interest included area-level SDOH, derived from the 2016 Canada Census at the level of dissemination areas (DAs), the smallest geographic unit (representing 400-700 residents) for which census data are reported (19). The list of area-level SDOH included factors reflecting SES (median household income, educational attainment, proportion essential workers), ethnic diversity (proportion racialized groups, proportion recent immigrants), and housing conditions (proportion apartment buildings, proportion high-density housing, average household size). Detailed definitions of these variables are shown in **footnotes of Table-1**.

**Table 1.**
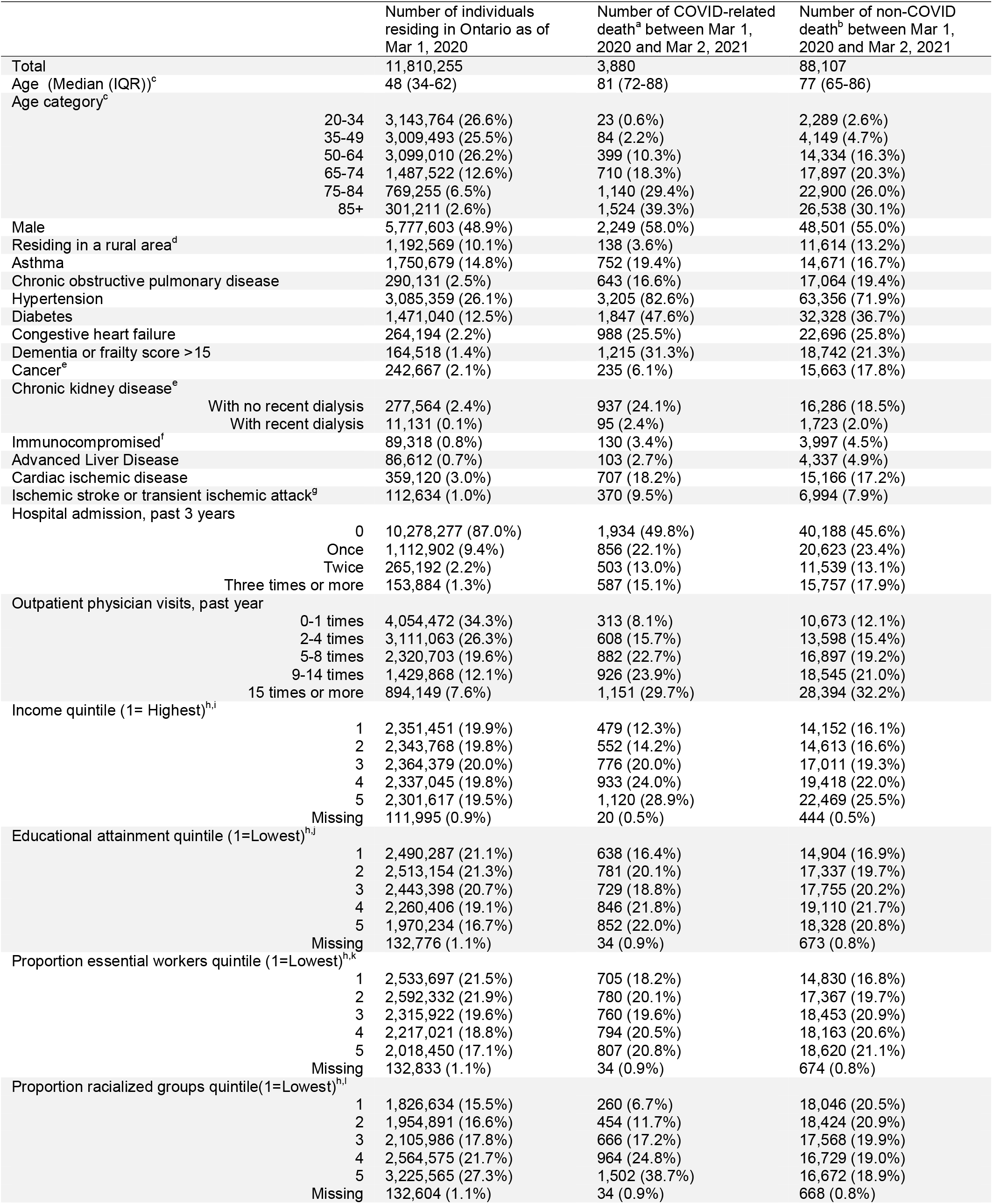
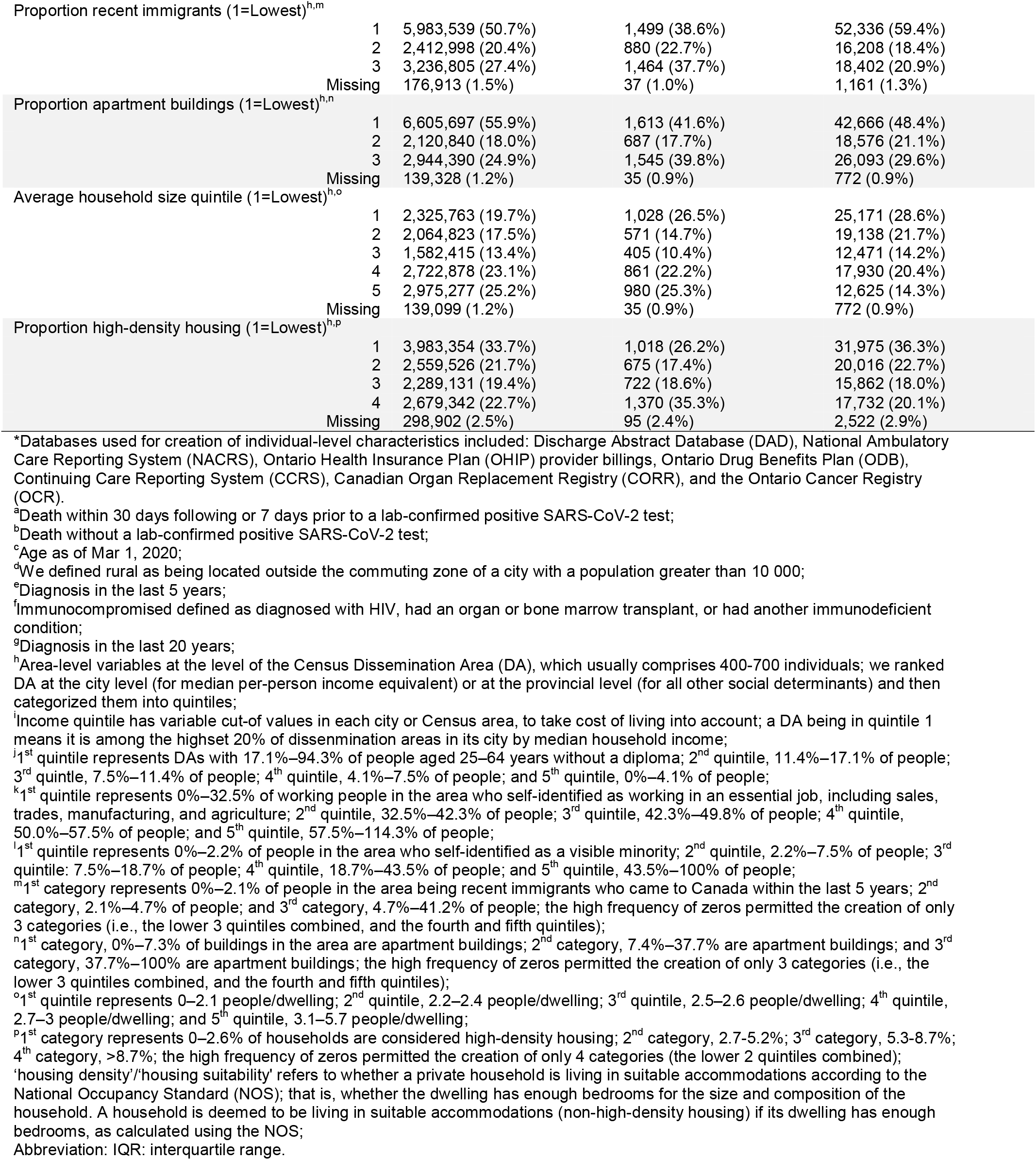
Characteristics* of overall community dwelling adults in Ontario and those died related to COVID-19 and other causes.

|  |  | Number of individuals<br>residing in Ontario as of<br>Mar 1, 2020 | Number of COVID-related<br>death<sup>a</sup> between Mar 1,<br>2020 and Mar 2, 2021 | Number of non-COVID<br>death<sup>b</sup> between Mar 1,<br>2020 and Mar 2, 2021 |
| --- | --- | --- | --- | --- |
| Total |  | 11,810,255 | 3,880 | 88,107 |
| Age (Median (IQR))<sup>c</sup> |  | 48 (34-62) | 81 (72-88) | 77 (65-86) |
| Age category<sup>c</sup> |  |  |  |  |
|  | 20-34 | 3,143,764 (26.6%) | 23 (0.6%) | 2,289 (2.6%) |
|  | 35-49 | 3,009,493 (25.5%) | 84 (2.2%) | 4,149 (4.7%) |
|  | 50-64 | 3,099,010 (26.2%) | 399 (10.3%) | 14,334 (16.3%) |
|  | 65-74 | 1,487,522 (12.6%) | 710 (18.3%) | 17,897 (20.3%) |
|  | 75-84 | 769,255 (6.5%) | 1,140 (29.4%) | 22,900 (26.0%) |
|  | 85+ | 301,211 (2.6%) | 1,524 (39.3%) | 26,538 (30.1%) |
| Male |  | 5,777,603 (48.9%) | 2,249 (58.0%) | 48,501 (55.0%) |
| Residing in a rural area<sup>d</sup> |  | 1,192,569 (10.1%) | 138 (3.6%) | 11,614 (13.2%) |
| Asthma |  | 1,750,679 (14.8%) | 752 (19.4%) | 14,671 (16.7%) |
| Chronic obstructive pulmonary disease |  | 290,131 (2.5%) | 643 (16.6%) | 17,064 (19.4%) |
| Hypertension |  | 3,085,359 (26.1%) | 3,205 (82.6%) | 63,356 (71.9%) |
| Diabetes |  | 1,471,040 (12.5%) | 1,847 (47.6%) | 32,328 (36.7%) |
| Congestive heart failure |  | 264,194 (2.2%) | 988 (25.5%) | 22,696 (25.8%) |
| Dementia or frailty score >15 |  | 164,518 (1.4%) | 1,215 (31.3%) | 18,742 (21.3%) |
| Cancer<sup>e</sup> |  | 242,667 (2.1%) | 235 (6.1%) | 15,663 (17.8%) |
| Chronic kidney disease<sup>e</sup> |  |  |  |  |
|  | With no recent dialysis | 277,564 (2.4%) | 937 (24.1%) | 16,286 (18.5%) |
|  | With recent dialysis | 11,131 (0.1%) | 95 (2.4%) | 1,723 (2.0%) |
| Immunocompromised<sup>f</sup> |  | 89,318 (0.8%) | 130 (3.4%) | 3,997 (4.5%) |
| Advanced Liver Disease |  | 86,612 (0.7%) | 103 (2.7%) | 4,337 (4.9%) |
| Cardiac ischemic disease |  | 359,120 (3.0%) | 707 (18.2%) | 15,166 (17.2%) |
| Ischemic stroke or transient ischemic attack<sup>g</sup> |  | 112,634 (1.0%) | 370 (9.5%) | 6,994 (7.9%) |
| Hospital admission, past 3 years |  |  |  |  |
|  | 0 | 10,278,277 (87.0%) | 1,934 (49.8%) | 40,188 (45.6%) |
|  | Once | 1,112,902 (9.4%) | 856 (22.1%) | 20,623 (23.4%) |
|  | Twice | 265,192 (2.2%) | 503 (13.0%) | 11,539 (13.1%) |
|  | Three times or more | 153,884 (1.3%) | 587 (15.1%) | 15,757 (17.9%) |
| Outpatient physician visits, past year |  |  |  |  |
|  | 0-1 times | 4,054,472 (34.3%) | 313 (8.1%) | 10,673 (12.1%) |
|  | 2-4 times | 3,111,063 (26.3%) | 608 (15.7%) | 13,598 (15.4%) |
|  | 5-8 times | 2,320,703 (19.6%) | 882 (22.7%) | 16,897 (19.2%) |
|  | 9-14 times | 1,429,868 (12.1%) | 926 (23.9%) | 18,545 (21.0%) |
|  | 15 times or more | 894,149 (7.6%) | 1,151 (29.7%) | 28,394 (32.2%) |
| Income quintile (1= Highest)<sup>h,i</sup> |  |  |  |  |
|  | 1 | 2,351,451 (19.9%) | 479 (12.3%) | 14,152 (16.1%) |
|  | 2 | 2,343,768 (19.8%) | 552 (14.2%) | 14,613 (16.6%) |
|  | 3 | 2,364,379 (20.0%) | 776 (20.0%) | 17,011 (19.3%) |
|  | 4 | 2,337,045 (19.8%) | 933 (24.0%) | 19,418 (22.0%) |
|  | 5 | 2,301,617 (19.5%) | 1,120 (28.9%) | 22,469 (25.5%) |
|  | Missing | 111,995 (0.9%) | 20 (0.5%) | 444 (0.5%) |
| Educational attainment quintile (1=Lowest)<sup>h,j</sup> |  |  |  |  |
|  | 1 | 2,490,287 (21.1%) | 638 (16.4%) | 14,904 (16.9%) |
|  | 2 | 2,513,154 (21.3%) | 781 (20.1%) | 17,337 (19.7%) |
|  | 3 | 2,443,398 (20.7%) | 729 (18.8%) | 17,755 (20.2%) |
|  | 4 | 2,260,406 (19.1%) | 846 (21.8%) | 19,110 (21.7%) |
|  | 5 | 1,970,234 (16.7%) | 852 (22.0%) | 18,328 (20.8%) |
|  | Missing | 132,776 (1.1%) | 34 (0.9%) | 673 (0.8%) |
| Proportion essential workers quintile (1=Lowest)<sup>h,k</sup> |  |  |  |  |
|  | 1 | 2,533,697 (21.5%) | 705 (18.2%) | 14,830 (16.8%) |
|  | 2 | 2,592,332 (21.9%) | 780 (20.1%) | 17,367 (19.7%) |
|  | 3 | 2,315,922 (19.6%) | 760 (19.6%) | 18,453 (20.9%) |
|  | 4 | 2,217,021 (18.8%) | 794 (20.5%) | 18,163 (20.6%) |
|  | 5 | 2,018,450 (17.1%) | 807 (20.8%) | 18,620 (21.1%) |
|  | Missing | 132,833 (1.1%) | 34 (0.9%) | 674 (0.8%) |
| Proportion racialized groups quintile(1=Lowest)<sup>h,l</sup> |  |  |  |  |
|  | 1 | 1,826,634 (15.5%) | 260 (6.7%) | 18,046 (20.5%) |
|  | 2 | 1,954,891 (16.6%) | 454 (11.7%) | 18,424 (20.9%) |
|  | 3 | 2,105,986 (17.8%) | 666 (17.2%) | 17,568 (19.9%) |
|  | 4 | 2,564,575 (21.7%) | 964 (24.8%) | 16,729 (19.0%) |
|  | 5 | 3,225,565 (27.3%) | 1,502 (38.7%) | 16,672 (18.9%) |
|  | Missing | 132,604 (1.1%) | 34 (0.9%) | 668 (0.8%) |
| Proportion recent immigrants (1=Lowest)<sup>h,m</sup> |  |  |  |  |
|  | 1 | 5,983,539 (50.7%) | 1,499 (38.6%) | 52,336 (59.4%) |
|  | 2 | 2,412,998 (20.4%) | 880 (22.7%) | 16,208 (18.4%) |
|  | 3 | 3,236,805 (27.4%) | 1,464 (37.7%) | 18,402 (20.9%) |
|  | Missing | 176,913 (1.5%) | 37 (1.0%) | 1,161 (1.3%) |
| Proportion apartment buildings (1=Lowest)<sup>h,n</sup> |  |  |  |  |
|  | 1 | 6,605,697 (55.9%) | 1,613 (41.6%) | 42,666 (48.4%) |
|  | 2 | 2,120,840 (18.0%) | 687 (17.7%) | 18,576 (21.1%) |
|  | 3 | 2,944,390 (24.9%) | 1,545 (39.8%) | 26,093 (29.6%) |
|  | Missing | 139,328 (1.2%) | 35 (0.9%) | 772 (0.9%) |
| Average household size quintile (1=Lowest)<sup>h,o</sup> |  |  |  |  |
|  | 1 | 2,325,763 (19.7%) | 1,028 (26.5%) | 25,171 (28.6%) |
|  | 2 | 2,064,823 (17.5%) | 571 (14.7%) | 19,138 (21.7%) |
|  | 3 | 1,582,415 (13.4%) | 405 (10.4%) | 12,471 (14.2%) |
|  | 4 | 2,722,878 (23.1%) | 861 (22.2%) | 17,930 (20.4%) |
|  | 5 | 2,975,277 (25.2%) | 980 (25.3%) | 12,625 (14.3%) |
|  | Missing | 139,099 (1.2%) | 35 (0.9%) | 772 (0.9%) |
| Proportion high-density housing (1=Lowest)<sup>h,p</sup> |  |  |  |  |
|  | 1 | 3,983,354 (33.7%) | 1,018 (26.2%) | 31,975 (36.3%) |
|  | 2 | 2,559,526 (21.7%) | 675 (17.4%) | 20,016 (22.7%) |
|  | 3 | 2,289,131 (19.4%) | 722 (18.6%) | 15,862 (18.0%) |
|  | 4 | 2,679,342 (22.7%) | 1,370 (35.3%) | 17,732 (20.1%) |
|  | Missing | 298,902 (2.5%) | 95 (2.4%) | 2,522 (2.9%) |

\*Databases used for creation of individual-level characteristics included: Discharge Abstract Database (DAD), National Ambulatory Care Reporting System (NACRS), Ontario Health Insurance Plan (OHIP) provider billings, Ontario Drug Benefits Plan (ODB), Continuing Care Reporting System (CCRS), Canadian Organ Replacement Registry (CORR), and the Ontario Cancer Registry (OCR).
<sup>a</sup>Death within 30 days following or 7 days prior to a lab-confirmed positive SARS-CoV-2 test;
<sup>b</sup>Death without a lab-confirmed positive SARS-CoV-2 test;
<sup>c</sup>Age as of Mar 1, 2020;
<sup>d</sup>We defined rural as being located outside the commuting zone of a city with a population greater than 10 000;
<sup>e</sup>Diagnosis in the last 5 years;
<sup>f</sup>Immunocompromised defined as diagnosed with HIV, had an organ or bone marrow transplant, or had another immunodeficient condition;
<sup>g</sup>Diagnosis in the last 20 years;
<sup>h</sup>Area-level variables at the level of the Census Dissemination Area (DA), which usually comprises 400-700 individuals; we ranked DA at the city level (for median per-person income equivalent) or at the provincial level (for all other social determinants) and then categorized them into quintiles;
<sup>i</sup>Income quintile has variable cut-of values in each city or Census area, to take cost of living into account; a DA being in quintile 1 means it is among the highest 20% of dissemination areas in its city by median household income;
<sup>j</sup>1<sup>st</sup> quintile represents DAs with 17.1%–94.3% of people aged 25–64 years without a diploma; 2<sup>nd</sup> quintile, 11.4%–17.1% of people; 3<sup>rd</sup> quintile, 7.5%–11.4% of people; 4<sup>th</sup> quintile, 4.1%–7.5% of people; and 5<sup>th</sup> quintile, 0%–4.1% of people;
<sup>k</sup>1<sup>st</sup> quintile represents 0%–32.5% of working people in the area who self-identified as working in an essential job, including sales, trades, manufacturing, and agriculture; 2<sup>nd</sup> quintile, 32.5%–42.3% of people; 3<sup>rd</sup> quintile, 42.3%–49.8% of people; 4<sup>th</sup> quintile, 50.0%–57.5% of people; and 5<sup>th</sup> quintile, 57.5%–114.3% of people;
<sup>l</sup>1<sup>st</sup> quintile represents 0%–2.2% of people in the area who self-identified as a visible minority; 2<sup>nd</sup> quintile, 2.2%–7.5% of people; 3<sup>rd</sup> quintile, 7.5%–18.7% of people; 4<sup>th</sup> quintile, 18.7%–43.5% of people; and 5<sup>th</sup> quintile, 43.5%–100% of people;
<sup>m</sup>1<sup>st</sup> category represents 0%–2.1% of people in the area being recent immigrants who came to Canada within the last 5 years; 2<sup>nd</sup> category, 2.1%–4.7% of people; and 3<sup>rd</sup> category, 4.7%–41.2% of people; the high frequency of zeros permitted the creation of only 3 categories (i.e., the lower 3 quintiles combined, and the fourth and fifth quintiles);

All covariates other than SDOH were measured at the individual-level. We obtained data using heath administrative databases and disease registries, on age and sex, other demographics (living in rural vs. urban, and public health region), and baseline health. Measures of baseline health included a set of comorbidity variables, hospital admission in the past three years, and outpatient physician visits in the past year.

All data sets were linked using unique encoded identifiers and analyzed at ICES.

### Statistical analysis

We examined and compared the demographics, baseline health, and SDOH of overall community-dwelling individuals, individuals who died related to a SARS-CoV-2 infection, and individuals who died without any prior SARS-CoV-2 infection using descriptive statistics.

To examine the relationship between SDOH and COVID-19-related mortality, we employed cause-specific hazard models(17, 25), where death without a positive SARS-CoV-2 test were treated as competing risk events, and censored at time of death (**Appendix-Figure-1**). We proposed *a priori* and fitted unadjusted, and a set of adjusted models with serial adjustment (adjusting for age and sex; adjusting for age, sex, plus other demographics; additionally adjusting for baseline health; and further adjusting for other SDOH) to assess the impact of different sets of confounders. The models were fitted using the PHREG procedure of SAS 9.4(26).

To compare patterns by SDOH in non-COVID-19 mortality to those in COVID-19-related mortality, we repeated the survival analyses using cause-specific hazard models to examine relationship between SDOH and non-COVID-19 mortality, treating COVID-19-related mortality as a competing risk.

To compare patterns by SDOH in COVID-19-related mortality to those in COVID-19 case fatality, we employed multivariable logistic regression models to examine the associations between SDOH and odds of COVID-19-related death among a subset of the study population who tested positive for SARS-CoV-2.

To quantify the absolute differences by SDOH in COVID-19-related mortality, we employed Fine & Gray subdistribution hazard models(17, 27). Based on the fitted models adjusted for demographics and baseline health, we estimated the adjusted marginal cumulative incidence functions(28) for each SDOH over a period of one year, and calculated the difference in the one-year cumulative probability of COVID-19-related death between the most and least vulnerable group of each SDOH variable.

## RESULTS

Of 11,810,255 community dwelling adults (median age 48 years, interquartile range: 34-62) included in our analysis, 206,671 (1.75%) tested positive for SARS-CoV-2, 3880 (0.03%) died within 30 days post to or 7 days prior to a positive test, and88,107 (0.75%) individuals died without a COVID-19 diagnosis.

Deaths related to a COVID-19 diagnosis were disproportionately concentrated among older adults, males and individuals living in urban areas(**Table-1**). COVID-19-related deaths were also disproportionately concentrated among individuals living with a comorbidity and with more prior healthcare use (**Table-1**). Compared to the overall community dwelling individuals, individuals who died related to a COVID-19 diagnosis were overrepresented in areas with less social advantage (e.g. 28.9% vs.19.5% lived in the lowest-income areas); and in areas with higher proportion of racialized groups (38.7% vs. 27.3%) and recent immigrants (37.7% vs. 27.4%) (**Table-1**).

### Associations between COVID-19-related mortality and area-level SDOH

In the unadjusted models, areas with lower SES, higher ethnic diversity, higher proportion apartment buildings and high-density housing, and the lowest or highest household size (vs. medium) were associated with increased hazard of COVID-19-related death (**Figure-2A, Appendix-Table-2**). We observed a dose-response relationship between all SDOH variables and COVID-19-related mortality, except household size (**Figure-2**).

**Figure 2.**
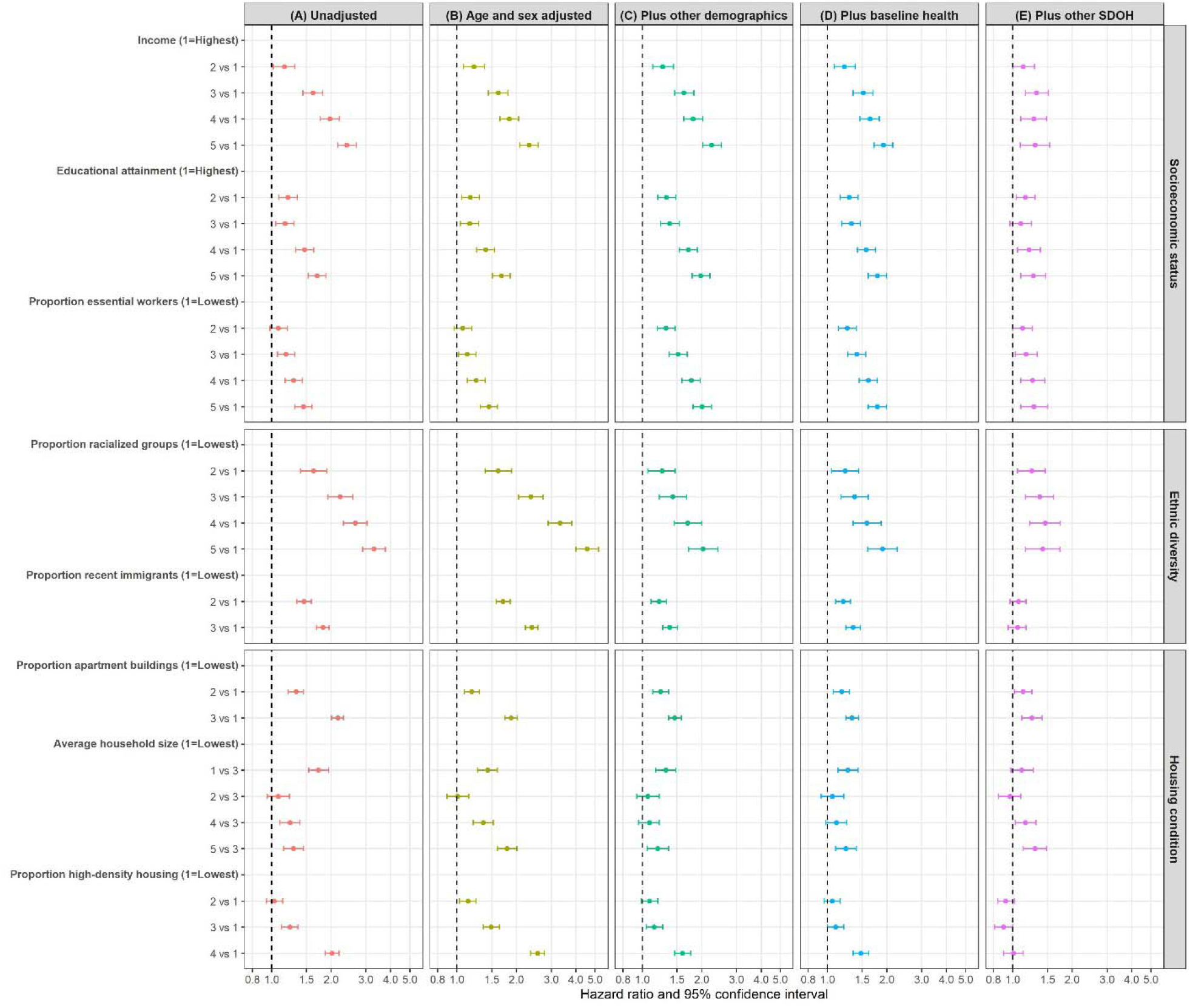
Associations between area-level social determinants of health (SDOH) and COVID-19-related mortality among community dwelling adult populations aged 20 years and older in Ontario, Canada between March 1, 2020 and Mar 2, 2021, with serial adjustment of potential confounders. Cause-specific hazard models were used for COVID-19-related mortality analyses. COVID-19-related death defined as death within 30 days following or 7 days prior to a positive SARS-CoV-2 test. Other demographics variables included whether individuals reside in rural vs. urban area, and the public health region where individuals reside. Baseline health variables included comorbidities (list in **Table 1**), number of hospital admissions in the past 3 years, and outpatient physician visits in the past year. Other SDOH variables are shown in the figure per Y-axis. All area-level SDOH variables are measured at the level of the Census Dissemination Area, except income (at census metropolitan area), and detailed definitions of these variables are shown in **Table 1** footnotes.

#### Socioeconomic status (SES): income, education, essential workers

Adjustment for demographics either attenuated or amplified the associations between COVID-19-related mortality and SES (**Figure-2A-2C**). Further adjustment for baseline health slightly reduced the associations between COVID-19 related mortality and SES (**Figure-2C-2D**). After further adjustment for other SDOH, SES remained an independent determinant of COVID-19 related mortality, although the magnitude of association was greatly reduced (**Figure-2D-2E**). Fully adjusted hazard ratios (aHRs) and 95% confidence intervals (CIs) were 1.30 [1.09,1.54] for lowest vs. highest income, 1.27 [1.10,1.47] for lowest vs. highest educational attainment, and 1.28 [1.10,1.50] for highest vs. lowest proportion essential workers (**Figure-2E and Appendix-Table-2**).

#### Ethnic diversity: racialized groups, recent immigrants

Adjustment for age and sex increased the magnitude of associations between ethnic diversity and the COVID-19-related mortality (**Figure-2A-2B**), while additional adjustment for other demographics largely reduced the magnitude of associations (**Figure-2B-2C**). Further adjustment for individual’s baseline health had a minimal influence on the associations (**Figure-2C-2D**). Additional adjustment of other SDOH reduced the magnitude of associations between COVID-19-related mortality and proportion racialized groups, and nullified the association between COVID-19-related mortality and proportion recent immigrants (**Figure-2D-2E**). Fully adjusted aHR and 95% CI were 1.42 [1.16,1.73] for highest vs. lowest proportion racialized groups (**Figure-2E and Appendix**-**Table-2**).

#### Housing condition: apartment buildings, high-density housing, household size

After adjustment for demographics and baseline health, and other SDOH, proportion apartment buildings was independently associated with increased hazard of COVID-19-related death (aHR and 95% CI were 1.25 [1.11,1.41]); while proportion high-density housing was not (**Figure-2E**; **Appendix-Table-2**). The non-monotonic relationship between COVID-19-related mortality and household size persisted after full adjustment. Fully adjusted aHR and 95% CI were 1.30 [1.13,1.48] for highest vs. medium household size (**Figure-2E and Appendix**-**Table-2**).

### Comparison of area-level SDOH in COVID-19-related mortality vs. non-COVID-19 mortality, and vs. COVID-19 case fatality

In contrast to the pattern with COVID-19-related mortality, areas with higher proportion racialized groups (highest vs. lowest: 0.88 [0.85,0.92]), and larger household size (highest vs medium: 0.85 [0.83,0.88]) were independently associated with decreased hazard of non-COVID-19 death (**Figure-3A-3B and Appendix-Table-3-4**).

**Figure 3.**
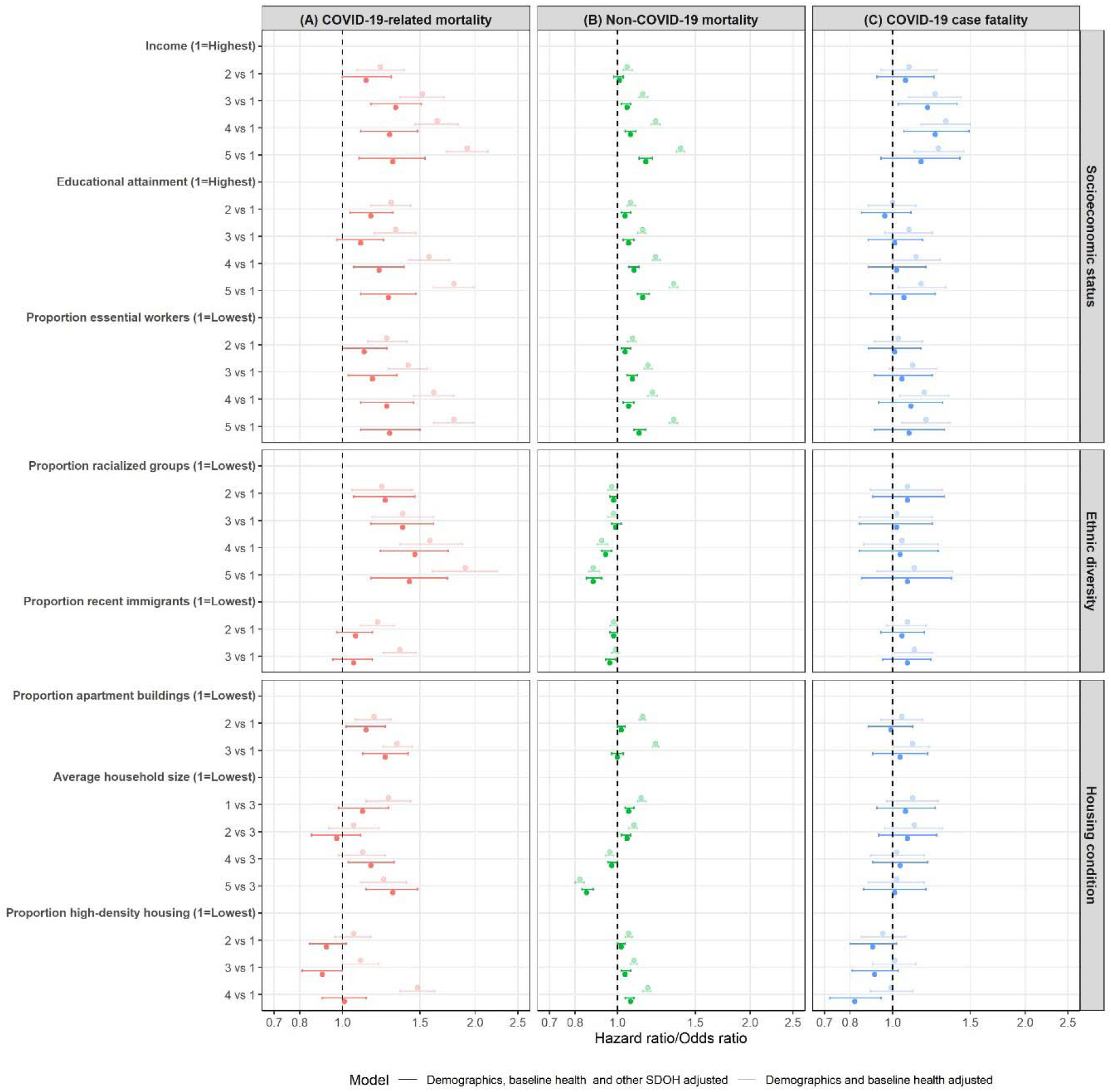
Comparing area-level social determinants of health (SDOH) in COVID-19-related mortality, non-COVID-19 mortality, and COVID-19 case fatality among community dwelling adult populations aged 20 years and older in Ontario, Canada, March 1 2020 – Mar 2, 2021. Multivariable cause-specific hazard models and logistic regression model were used to estimate cause-specific mortalities and case fatality, respectively. Death within 30 days following or 7 days prior to a positive SARS-CoV-2 test was considered in calculations of COVID-19 case fatality and COVID-19-related mortality. Death without a positive SARS-CoV-2 test was considered non-COVID-19 mortality. Demographics variables included age, sex, whether individuals reside in rural vs. urban area, and the public health region where individuals reside. Baseline health variables included comorbidities (list in **Table 1**), number of hospital admissions in the past 3 years, and outpatient physician visits in the past year. Other SDOH variables are shown per Y-axis. All area-level SDOH variables are measured at Census Dissemination Area level except income (at census metropolitan area), and detailed definitions of these variables are shown in **Table 1** footnotes. The case fatality model additionally adjusted for month of COVID-19 test.

In the fully adjusted model, only lower income was independently associated with increased COVID-19 case fatality (**Figure-3C and Appendix-Table-3-4**).

### Adjusted cumulative incidence of COVID-19-related death by SDOH

After accounting for demographics and baseline health, the estimated absolute difference in COVID-19-related death per 100,000 adults over a one-year period ranged from 6.4 to 20.3, comparing the most and least vulnerable SDOH group (**Figure-4**).

**Figure 4.**
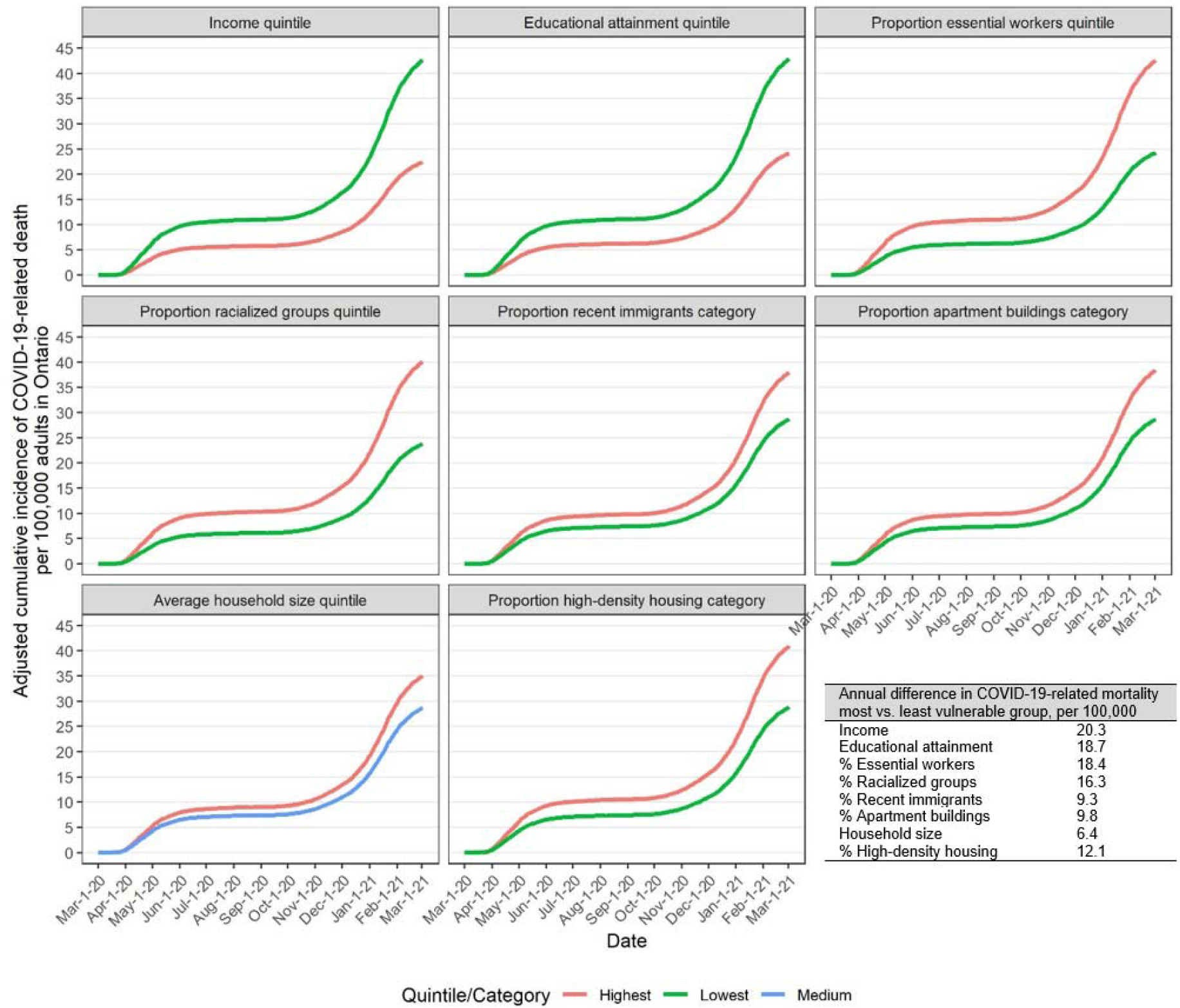
Adjusted cumulative incidence function of COVID-19-related mortality by area-level social determinants of health (SDOH) among community dwelling adult populations aged 20 years and older in Ontario, Canada, March 1 2020 – Mar 2, 2021. Death within 30 days following or 7 days prior to a positive SARS-CoV-2 test was considered COVID-19-related. Estimates were obtained from the fitted Fine & Gray subdistribution hazard models. The models adjusted for demographics (age, sex, whether individuals reside in rural vs. urban area, the public health region where individuals reside), and baseline health (comorbidities (list in **Table 1**), number of hospital admissions in the past 3 years, and outpatient physician visits in the past year). All area-level SDOH variables are measured at Census Dissemination Area level except income (at census metropolitan area), and detailed definitions of these variables are shown in **Table 1** footnotes.

## DISCUSSION

In a population-based cohort of 11.8 million adults in Ontario, Canada, we found that areas characterized by lower SES, greater ethnic diversity, more apartment buildings, and larger vs. medium household size were associated with increased hazard of COVID-19-related mortality, after accounting for individual-level demographics, baseline health, and other area-level SDOH. In contrast, areas with higher proportion racialized groups and larger household size were associated with reduced hazard of non-COVID-19 mortality. With the exception of income, the SDOH examined in this study were not independently associated with COVID-19 case fatality.

Our findings mirror studies in other countries, including the UK(24), Switzerland(13), Chile(9), and the US(14), which have shown that areas with lower SES, measured by a composite social vulnerability or deprivation index, were associated with increased risk and mortality of COVID-19. Our study demonstrated that specific elements of area-level SES, including income, educational attainment, and essential workers were each independently associated with elevated hazard of COVID-19-related mortality. For example, individuals working in front-facing essential services that were not amenable to remote work had limited ability to shelter-in-place during periods of broad-scale restrictions on mobility, and were less likely to receive benefits such as paid sick leave(29, 30), leading to heightened exposure risk and barriers to effective quarantine or isolation(8, 10). The relationship between income and case-fatality might reflect delayed diagnosis or access to and quality of clinical care for persons living in lower income neighbourhoods. Emerging evidence, including in Ontario, suggests that in-hospital mortality with COVID-19 is amplified during periods of higher patient load; such inpatient surges were most likely to also occur in hospitals serving lower income areas experiencing the highest rates of cases(31-34).

Our findings that areas with a higher proportion of racialized groups experienced increased hazard of COVID-19-related mortality, but not higher case-fatality confirmed findings in other settings(6, 15). A systematic review of 52 studies in the US found that African-American/Black and non-white Hispanic populations experienced a disproportionate burden of infections, hospitalization, and COVID-19-related mortality, but not higher in-hospital case-fatality, compared to similarly aged white non-Hispanic populations(6). Studies in the UK found that minority ethnic groups experienced elevated risk of COVID-19-related mortality(15), higher prevalence of SARS-CoV-2 antibodies(35), but similar infection fatality ratio(35) compared to white counterparts. Taken together, the findings suggest that inequalities in COVID-19-related mortality by racialized groups are more likely to stem from disproportionate exposure risks leading to disproportionate risks of acquisition and transmission, and barriers to the reach of, and access to prevention interventions; as opposed to differences post-diagnosis (6, 8, 15).

In Canada, racialized groups are more likely to work in essential services and more likely to live in larger and higher-density households(36)– all of which have been identified as mechanistic risk factors for heightened exposure risk(8, 10). Prior to COVID-19 and similar to our findings regarding non-COVID-19 mortality during the COVID-19 pandemic, mortality rates in Canada were lower in racialized groups(37). Similar to findings from the UK and Sweden (15, 38), COVID-19 has reversed the dose-response pattern of lower mortality by proportion racialized groups.

The non-monotonic relationship between household size and COVID-19-related mortality might be partially explained by the positive correlation between income and household size (data not shown). Despite potential residual confounding by income, our findings revealed that large household size, regardless of the housing density, might be an independent risk factor for household transmission. In epidemic theory, contact rates are conceptualized as density-dependent or frequency-dependent. Transmissions outside households may be influenced by population density (density-dependent transmission)(39). Within the same household, contact rates may be better reflected by the frequency-dependent transmission (i.e., assumes close interactions among all household members, regardless of the household density)(39). As such, while factors such as crowded spaces have been identified as important risk factors of transmission(8, 23), households may confer a different pattern wherein the number of household contacts (thus, household size) may be just as, or potentially more important, than housing density alone for within-household transmission.

Strengths of our study include limiting selection bias; and leveraging high-quality linked health administrative, surveillance, and health registries data to examine the influence of various confounders, including comorbidities on the relationship between COVID-19-related mortality and SDOH. Another strength is the competing risk survival analysis approach which allowed us to correctly estimate the marginal probability of COVID-19-related death in the presence of competing events. Our estimates of marginal probability of COVID-19-related death by SDOH provided important insights into health of each subgroup, beyond the health equality itself, and permitted the quantification of adjusted inequalities on an absolute scale, in addition to the relative scale widely used in the literature(13, 15, 40). Measures of inequalities on an absolute scale provided a tangible quantification of the impact of inequalities, which are meaningful for public health decision making including informing strategies such as geographically-focused vaccination (41-43).

Limitations include the potential for misclassification biases due to lack of data on the cause of death. Based on the Ontario COIVD-19 surveillance data, 92% of recorded all-cause death among individuals diagnosed with COVID-19 occurred within 30 days following or 7 days prior to a positive test (**Appendix-Figure-2**). Other settings have adopted similar definition of COVID-19-related death to capture the immediate impact of COVID-19 on death(44). Individuals who do not have a health card number were not captured; and if they were more likely to be socially and structurally vulnerable, our estimates might have under-estimated the inequalities. We were restricted to area-level SDOH measures in the absence of individual-level SDOH measures, leading to the potential for residual confounders by SDOH. However, our findings were similar in pattern to that of other studies using individual-level SDOH measures(3, 15), suggesting a lower risk of ecological fallacy. Almost all areas with the highest quintile proportion racialized groups were urban areas. Therefore, our estimated associations between racialized groups and COVID-19-related mortality are subject to residual confounding by individual’s residence in rural vs urban areas. However, stratified analysis by rural/urban revealed that structural inequalities in COVID-19-related mortality by racialized groups were present in both settings (**Appendix-Table-5**). We did not examine the potential modifying effect by age or region, nor the potential changes over time (e.g., between pandemic waves, or in the context of vaccination) in the magnitude of inequalities(9, 15); which will be an important next step of research.

Our study demonstrated that area-level social and structural inequalities determine COVID-19-related mortality even after accounting for age, sex, and clinical factors. The majority of inequalities stem from proximal exposures and reach of, and access to prevention interventions. COVID-19 has reversed existing pattern of mortality by race/ethnicity, with higher COVID-19-related mortality for racialized groups. Tailored strategies that specifically address and are designed around the risk pathways related to SES, systemic racism, and housing context, include but are not limited to: paid sick leave and improved workplace health and safety protocols and outbreak management; and community-led and community-tailored outreach for testing, effective isolation and quarantine and vaccine programs. Moving forward, the goal of pandemic responses should include improving overall health of population via addressing the SDOH that mechanistically underpin social and structural inequalities in acquisition and transmission risks, and shape barriers to the reach of, and access to prevention interventions.

## Data Availability

The data set from this study is held securely in coded form at ICES. While legal data-sharing agreements between ICES and data providers (e.g., health care organizations and government) prohibit ICES from making the data set publicly available, access may be granted to those who meet prespecified criteria for confidential access, available at www.ices.on.ca/DAS. The full dataset creation plan and underlying analytic code are available from the authors upon request, understanding that the computer programs may rely upon coding templates or macros that are unique to ICES and are therefore either inaccessible or may require modification.

## Contribution

LW, JCK, and SM conceptualized the study. AC conducted the data cleaning and statistical analyses. LW drafted the manuscript. AC, SB, JS, AKC, BS, PCA, JCK, and SM provided critical inputs into the results interpretation and manuscript.

## Acknowledgements and disclaimers

The authors thank IQVIA Solutions Canada for use of their Drug Information File. The authors are grateful to the 14.7 million Ontario residents without whom this research would be impossible.

Parts of this material are based on data and/or information compiled and provided by the Canadian Institute for Health Information (CIHI) and by Cancer Care Ontario (CCO). However, the analyses, conclusions, opinions, and statement expressed herein are those of the authors, and not necessarily those of CIHI or CCO. This study was supported by ICES, which is funded by an annual grant from the Ontario Ministry of Health (MOH) and Ministry of Long-Term Care (MLTC). The study sponsors did not participate in the design and conduct of the study; collection, management, analysis and interpretation of the data; preparation, review or approval of the manuscript or the decision to submit the manuscript for publication.

The opinions, results, and conclusions reported in this paper are those of the authors and are independent from the funding sources. No endorsement by ICES, MOH, MLTC, CIHI or CCO is intended or should be inferred.

