## Appendix for "Area-level social and structural inequalities determine mortality related to COVID-19 diagnosis in Ontario, Canada: a population-based explanatory modeling study of 11.8 million people"

**Appendix Table 1. Completeness of COVID-19-related death**^a^ **records captured by CCM and RPDB**^b^**.**

| Year-month of death | Among COVID-19-related deaths captured by CCM, % missed by RPDB | Among COVID-19 related deaths captured by RPDB, % missed by CCM | Estimated^c^ % COVID-19-related deaths missed using both CCM and RPDB |
| --- | --- | --- | --- |
| 2020-03 | 0.0 | 1.7 | 0.0 |
| 2020-04 | 2.2 | 2.2 | 0.05 |
| 2020-05 | 1.0 | 5.4 | 0.05 |
| 2020-06 | 1.4 | 10.1 | 0.14 |
| 2020-07 | 0.0 | 10.3 | 0.0 |
| 2020-08 | 0.0 | 0.0 | 0.0 |
| 2020-09 | 3.8 | 7.4 | 0.28 |
| 2020-10 | 2.6 | 14.8 | 0.38 |
| 2020-11 | 2.0 | 19.1 | 0.38 |
| 2020-12 | 1.8 | 20.1 | 0.36 |
| 2021-01 | 3.9 | 24.6 | 0.96 |
| 2021-02 | 9.3 | 28.3 | 2.64 |
| 2021-03 | 11.5 | 29.2 | 3.35 |
| TOTAL | 3.7 | 18.6 | 0.69 |

^a^Death within 30 days following or 7 days prior to a lab-confirmed positive SARS-CoV-2 test;

^b^We used the CCM dataset and RPDB dataset updated as of July 5, 2021; we used death records in RPDB to supplement deaths recorded in CCM, because not all individuals who died due to COVID-19 were captured by the surveillance system due to limited capacity during peaks of outbreak;

^c^Estimated by (Among COVID-19-related deaths captured by CCM, % missed by RPDB) * (Among COVID-19-related deaths captured by RPDB, % missed by CCM);

Abbreviations: CCM, COVID-19 Case and Contact Management System; RPDB, Registered Persons Database.

**Appendix Table 2.** Associations between area-level social determinants of health (SDOH) and COVID-19-related mortality^a^ among community dwelling adult populations aged 20 years and older in Ontario, Canada between March 1, 2020 and Mar 2, 2021, with serial adjustment of potential confounders.

|  | Unadjusted | Age and sex adjusted | Plus other demograhics^b^ | Plus baseline health^c^ | Plus other SDOH^d^ |
| --- | --- | --- | --- | --- | --- |
| Area-level SDOH^d^ | Hazard ratio^e^(95% confidence interval) | | | | |
| Income (1= Highest) |  |  |  |  |  |
| 2 vs 1 | 1.16 (1.02, 1.31) | 1.22 (1.08, 1.38) | 1.27 (1.13, 1.44) | 1.22 (1.08, 1.38) | 1.13 (1.00, 1.29) |
| 3 vs 1 | 1.62 (1.44, 1.81) | 1.62 (1.44, 1.81) | 1.63 (1.46, 1.83) | 1.52 (1.35, 1.70) | 1.32 (1.16, 1.51) |
| 4 vs 1 | 1.97 (1.76, 2.20) | 1.84 (1.65, 2.05) | 1.81 (1.62, 2.02) | 1.64 (1.46, 1.83) | 1.28 (1.10, 1.48) |
| 5 vs 1 | 2.40 (2.16, 2.68) | 2.32 (2.08, 2.58) | 2.25 (2.02, 2.51) | 1.92 (1.72, 2.14) | 1.30 (1.09, 1.54) |
| Educational attainment (1=Highest) |  |  |  |  |  |
| 2 vs 1 | 1.21 (1.09, 1.35) | 1.17 (1.06, 1.30) | 1.33 (1.20, 1.48) | 1.29 (1.16, 1.43) | 1.16 (1.04, 1.30) |
| 3 vs 1 | 1.17 (1.05, 1.30) | 1.16 (1.04, 1.29) | 1.38 (1.24, 1.54) | 1.32 (1.18, 1.47) | 1.10 (0.97, 1.24) |
| 4 vs 1 | 1.47 (1.32, 1.63) | 1.40 (1.26, 1.55) | 1.71 (1.54, 1.90) | 1.57 (1.42, 1.75) | 1.21 (1.06, 1.38) |
| 5 vs 1 | 1.70 (1.53, 1.88) | 1.68 (1.51, 1.86) | 1.98 (1.79, 2.20) | 1.79 (1.61, 1.99) | 1.27 (1.10, 1.47) |
| Proportion essential workers (1=Lowest) |  |  |  |  |  |
| 2 vs 1 | 1.08 (0.98, 1.20) | 1.07 (0.97, 1.19) | 1.32 (1.19, 1.47) | 1.26 (1.14, 1.40) | 1.12 (1.00, 1.26) |
| 3 vs 1 | 1.18 (1.07, 1.31) | 1.13 (1.02, 1.25) | 1.52 (1.37, 1.69) | 1.41 (1.27, 1.56) | 1.17 (1.03, 1.33) |
| 4 vs 1 | 1.29 (1.17, 1.43) | 1.25 (1.13, 1.39) | 1.77 (1.59, 1.97) | 1.61 (1.45, 1.79) | 1.26 (1.10, 1.45) |
| 5 vs 1 | 1.45 (1.31, 1.60) | 1.45 (1.31, 1.60) | 2.01 (1.81, 2.24) | 1.79 (1.61, 1.99) | 1.28 (1.10, 1.50) |
| Proportion racialized groups (1=Lowest) |  |  |  |  |  |
| 2 vs 1 | 1.63 (1.40, 1.90) | 1.62 (1.39, 1.89) | 1.26 (1.07, 1.47) | 1.23 (1.05, 1.44) | 1.25 (1.06, 1.46) |
| 3 vs 1 | 2.22 (1.93, 2.57) | 2.37 (2.05, 2.73) | 1.43 (1.22, 1.68) | 1.37 (1.17, 1.61) | 1.37 (1.16, 1.61) |
| 4 vs 1 | 2.65 (2.31, 3.04) | 3.32 (2.89, 3.81) | 1.70 (1.45, 2.00) | 1.58 (1.35, 1.87) | 1.46 (1.22, 1.74) |
| 5 vs 1 | 3.30 (2.89, 3.76) | 4.56 (4.00, 5.21) | 2.03 (1.72, 2.41) | 1.90 (1.60, 2.25) | 1.42 (1.16, 1.73) |
| Proportion recent immigrants (1=Lowest) |  |  |  |  |  |
| 2 vs 1 | 1.46 (1.34, 1.59) | 1.71 (1.58, 1.86) | 1.22 (1.11, 1.33) | 1.20 (1.10, 1.31) | 1.07 (0.97, 1.17) |
| 3 vs 1 | 1.82 (1.69, 1.95) | 2.39 (2.22, 2.57) | 1.38 (1.27, 1.51) | 1.35 (1.24, 1.47) | 1.06 (0.95, 1.17) |
| Proportion apartment buildings (1=Lowest) |  |  |  |  |  |
| 2 vs 1 | 1.33 (1.21, 1.45) | 1.19 (1.09, 1.30) | 1.24 (1.13, 1.36) | 1.18 (1.07, 1.29) | 1.13 (1.02, 1.25) |
| 3 vs 1 | 2.16 (2.01, 2.31) | 1.88 (1.75, 2.02) | 1.46 (1.36, 1.58) | 1.33 (1.24, 1.44) | 1.25 (1.11, 1.41) |
| Average household size (1=Lowest) |  |  |  |  |  |
| 1 vs 3 | 1.73 (1.54, 1.94) | 1.43 (1.27, 1.60) | 1.32 (1.17, 1.48) | 1.27 (1.13, 1.43) | 1.11 (0.98, 1.27) |
| 2 vs 3 | 1.08 (0.95, 1.23) | 1.01 (0.89, 1.15) | 1.07 (0.94, 1.22) | 1.06 (0.93, 1.21) | 0.97 (0.85, 1.10) |
| 4 vs 3 | 1.24 (1.10, 1.39) | 1.36 (1.21, 1.53) | 1.09 (0.96, 1.22) | 1.11 (0.98, 1.25) | 1.16 (1.03, 1.31) |
| 5 vs 3 | 1.29 (1.15, 1.45) | 1.79 (1.60, 2.01) | 1.20 (1.06, 1.36) | 1.24 (1.10, 1.40) | 1.30 (1.13, 1.48) |
| Proportion high-density housing (1=Lowest) |  |  |  |  |  |
| 2 vs 1 | 1.03 (0.94, 1.14) | 1.14 (1.03, 1.25) | 1.09 (0.99, 1.20) | 1.06 (0.96, 1.16) | 0.92 (0.84, 1.02) |
| 3 vs 1 | 1.24 (1.12, 1.36) | 1.49 (1.36, 1.64) | 1.15 (1.05, 1.27) | 1.10 (1.00, 1.21) | 0.90 (0.81, 1.00) |
| 4 vs 1 | 2.02 (1.86, 2.19) | 2.56 (2.36, 2.77) | 1.60 (1.46, 1.76) | 1.48 (1.35, 1.62) | 1.01 (0.90, 1.13) |

^a^Death within 30 days following or 7 days prior to a lab-confirmed positive SARS-CoV-2 test was considered COVID-19-related;

^b^Other demographics variables included whether individuals reside in rural vs. urban area, and the public health region where individuals reside;

^c^Baseline health variables included comorbidities (list in **Table 1**), number of hospital admissions in the past 3 years, and outpatient physician visits in the past year;

^d^All area-level SDOH variables are measured at the level of the Census Dissemination Area, and detailed definitions of these variables are shown in **Table 1** footnotes;

^e^Cause-specific hazard models were used for COVID-19-related mortality analyses.

**Appendix Table 3.** Comparing area-level social determinants of health (SDOH) in COVID-19-related mortality^a^, non-COVID-19 mortality^b^, and COVID-19 case fatality^c^ among community dwelling adult populations aged 20 years and older in Ontario, Canada, March 1 2020 – Mar 2, 2021.

|  | COVID-19-related mortality | | Non-COVID-19 mortality | | COVID-19 case fatality | |
| --- | --- | --- | --- | --- | --- | --- |
| Area-level SDOH^d^ | Hazard ratio^e^ (95% CI) | | Hazard ratio^e^(95% CI) | | Odds ratio^e^ (95% CI) | |
|  | Partially adjusted^f^ | Fully adjusted^f^ | Partially adjusted^f^ | Fully adjusted^f^ | Partially adjusted^f^ | Fully adjusted^f^ |
| Income (1= Highest) |  |  |  |  |  |  |
| 2 vs 1 | 1.22 (1.08, 1.38) | 1.13 (1.00, 1.29) | 1.05 (1.03, 1.08) | 1.01 (0.98, 1.03) | 1.09 (0.94, 1.26) | 1.07 (0.92, 1.24) |
| 3 vs 1 | 1.52 (1.35, 1.70) | 1.32 (1.16, 1.51) | 1.14 (1.12, 1.17) | 1.05 (1.02, 1.07) | 1.25 (1.09, 1.43) | 1.20 (1.03, 1.40) |
| 4 vs 1 | 1.64 (1.46, 1.83) | 1.28 (1.10, 1.48) | 1.22 (1.19, 1.25) | 1.07 (1.04, 1.10) | 1.32 (1.16, 1.50) | 1.25 (1.06, 1.49) |
| 5 vs 1 | 1.92 (1.72, 2.14) | 1.30 (1.09, 1.54) | 1.39 (1.36, 1.42) | 1.16 (1.12, 1.20) | 1.27 (1.12, 1.45) | 1.16 (0.94, 1.42) |
| Educational attainment (1=Highest) |  |  |  |  |  |  |
| 2 vs 1 | 1.29 (1.16, 1.43) | 1.16 (1.04, 1.30) | 1.07 (1.05, 1.10) | 1.04 (1.02, 1.07) | 1.00 (0.88, 1.13) | 0.96 (0.85, 1.10) |
| 3 vs 1 | 1.32 (1.18, 1.47) | 1.10 (0.97, 1.24) | 1.14 (1.11, 1.16) | 1.06 (1.03, 1.09) | 1.09 (0.96, 1.23) | 1.01 (0.88, 1.17) |
| 4 vs 1 | 1.57 (1.42, 1.75) | 1.21 (1.06, 1.38) | 1.22 (1.20, 1.25) | 1.09 (1.06, 1.12) | 1.13 (1.00, 1.28) | 1.02 (0.88, 1.19) |
| 5 vs 1 | 1.79 (1.61, 1.99) | 1.27 (1.10, 1.47) | 1.34 (1.31, 1.37) | 1.14 (1.11, 1.18) | 1.16 (1.03, 1.32) | 1.06 (0.89, 1.25) |
| Proportion essential workers (1=Lowest) |  |  |  |  |  |  |
| 2 vs 1 | 1.26 (1.14, 1.40) | 1.12 (1.00, 1.26) | 1.08 (1.05, 1.10) | 1.04 (1.02, 1.07) | 1.03 (0.91, 1.17) | 1.01 (0.88, 1.16) |
| 3 vs 1 | 1.41 (1.27, 1.56) | 1.17 (1.03, 1.33) | 1.17 (1.15, 1.20) | 1.08 (1.05, 1.11) | 1.11 (0.98, 1.26) | 1.05 (0.91, 1.23) |
| 4 vs 1 | 1.61 (1.45, 1.79) | 1.26 (1.10, 1.45) | 1.20 (1.17, 1.23) | 1.06 (1.03, 1.09) | 1.18 (1.04, 1.34) | 1.10 (0.93, 1.30) |
| 5 vs 1 | 1.79 (1.61, 1.99) | 1.28 (1.10, 1.50) | 1.34 (1.31, 1.37) | 1.12 (1.09, 1.16) | 1.19 (1.05, 1.35) | 1.09 (0.91, 1.31) |
| Proportion racialized groups (1=Lowest) |  |  |  |  |  |  |
| 2 vs 1 | 1.23 (1.05, 1.44) | 1.25 (1.06, 1.46) | 0.97 (0.95, 1.00) | 0.98 (0.96, 1.00) | 1.08 (0.89, 1.30) | 1.08 (0.90, 1.31) |
| 3 vs 1 | 1.37 (1.17, 1.61) | 1.37 (1.16, 1.61) | 0.98 (0.95, 1.00) | 0.99 (0.97, 1.02) | 1.02 (0.84, 1.23) | 1.02 (0.84, 1.23) |
| 4 vs 1 | 1.58 (1.35, 1.87) | 1.46 (1.22, 1.74) | 0.92 (0.90, 0.95) | 0.94 (0.92, 0.97) | 1.05 (0.86, 1.27) | 1.04 (0.84, 1.27) |
| 5 vs 1 | 1.90 (1.60, 2.25) | 1.42 (1.16, 1.73) | 0.88 (0.86, 0.91) | 0.88 (0.85, 0.92) | 1.12 (0.92, 1.37) | 1.08 (0.85, 1.36) |
| Proportion recent immigrants (1=Lowest) |  |  |  |  |  |  |
| 2 vs 1 | 1.20 (1.10, 1.31) | 1.07 (0.97, 1.17) | 0.98 (0.96, 1.00) | 0.98 (0.96, 1.00) | 1.08 (0.97, 1.19) | 1.05 (0.94, 1.18) |
| 3 vs 1 | 1.35 (1.24, 1.47) | 1.06 (0.95, 1.17) | 0.99 (0.97, 1.01) | 0.96 (0.94, 0.99) | 1.12 (1.01, 1.23) | 1.08 (0.95, 1.22) |
| Proportion apartment buildings (1=Lowest) |  |  |  |  |  |  |
| 2 vs 1 | 1.18 (1.07, 1.29) | 1.13 (1.02, 1.25) | 1.14 (1.12, 1.16) | 1.02 (1.00, 1.04) | 1.05 (0.94, 1.17) | 0.99 (0.88, 1.11) |
| 3 vs 1 | 1.33 (1.24, 1.44) | 1.25 (1.11, 1.41) | 1.22 (1.20, 1.24) | 1.00 (0.97, 1.03) | 1.11 (1.01, 1.21) | 1.04 (0.90, 1.20) |
| Average household size (1=Lowest) |  |  |  |  |  |  |
| 1 vs 3 | 1.27 (1.13, 1.43) | 1.11 (0.98, 1.27) | 1.13 (1.11, 1.16) | 1.06 (1.04, 1.09) | 1.11 (0.97, 1.27) | 1.07 (0.92, 1.25) |
| 2 vs 3 | 1.06 (0.93, 1.21) | 0.97 (0.85, 1.10) | 1.09 (1.06, 1.11) | 1.05 (1.02, 1.07) | 1.12 (0.96, 1.30) | 1.08 (0.93, 1.26) |
| 4 vs 3 | 1.11 (0.98, 1.25) | 1.16 (1.03, 1.31) | 0.96 (0.94, 0.98) | 0.97 (0.95, 1.00) | 1.02 (0.89, 1.18) | 1.04 (0.90, 1.20) |
| 5 vs 3 | 1.24 (1.10, 1.40) | 1.30 (1.13, 1.48) | 0.82 (0.80, 0.84) | 0.85 (0.83, 0.88) | 1.02 (0.88, 1.18) | 1.01 (0.86, 1.19) |
| Proportion high-density housing (1=Lowest) |  |  |  |  |  |  |
| 2 vs 1 | 1.06 (0.96, 1.16) | 0.92 (0.84, 1.02) | 1.06 (1.04, 1.08) | 1.02 (1.00, 1.04) | 0.95 (0.85, 1.07) | 0.90 (0.80, 1.02) |
| 3 vs 1 | 1.10 (1.00, 1.21) | 0.90 (0.81, 1.00) | 1.09 (1.07, 1.11) | 1.04 (1.02, 1.07) | 1.01 (0.90, 1.13) | 0.91 (0.81, 1.03) |
| 4 vs 1 | 1.48 (1.35, 1.62) | 1.01 (0.90, 1.13) | 1.17 (1.14, 1.19) | 1.07 (1.04, 1.09) | 0.99 (0.89, 1.11) | 0.82 (0.72, 0.94) |

^a^Death within 30 days following or 7 days prior to a lab-confirmed positive SARS-CoV-2 test was considered COVID-19-related; ^b^Death without a lab-confirmed positive SARS-CoV-2 test; ^c^Death within 30 days following or 7 days prior to a lab-confirmed positive SARS-CoV-2 test among those diagnosed with COVID-19;

^d^All area-level SDOH variables are measured at the level of the Census Dissemination Area, and detailed definitions of these variables are shown in **Table 1** footnotes;

^e^Cause-specific hazard models were used for COVID-19-related mortality analyses, and non-COVID-19 mortality analyses; and logistic regression models were used for case fatality analyses; ^f^partially adjusted models adjusted for demographics (age, sex, whether individuals reside in rural vs. urban area, the public health region where individuals reside), and baseline health (comorbidities (list in **Table 1**), number of hospital admissions in the past 3 years, and outpatient physician visits in the past year); fully adjusted models additionally adjusted for the list of area-level SDOH shown in the table; both the partially and fully adjusted case fatality models additionally adjusted for month of COVID-19 test;

Abbreviations: CI, confidence interval.

**Appendix Table 4.** Comparing independent determinants for COVID-19-related mortality^a^, non-COVID-19 mortality^b^, and COVID-19 case fatality^c^ among community dwelling adult populations aged 20 years and older in Ontario, Canada, March 1 2020 – Mar 2, 2021.

|  | COVID-19-related mortality | Non-COVID-19 mortality | COVID-19 case fatality |
| --- | --- | --- | --- |
|  | Hazard ratio^d^(95% CI) | | Odds ratio^d^ (95% CI) |
| Age category |  |  |  |
| 20-24 vs 40-45 | 0.15 (0.05, 0.50) | 0.47 (0.42, 0.52) | 0.09 (0.02, 0.38) |
| 25-29 vs 40-45 | 0.40 (0.19, 0.84) | 0.58 (0.53, 0.63) | 0.26 (0.12, 0.59) |
| 30-34 vs 40-45 | 0.37 (0.18, 0.78) | 0.64 (0.59, 0.70) | 0.33 (0.15, 0.71) |
| 35-39 vs 40-45 | 0.58 (0.31, 1.11) | 0.78 (0.72, 0.84) | 0.52 (0.27, 1.02) |
| 45-49 vs 40-45 | 1.78 (1.08, 2.92) | 1.29 (1.20, 1.38) | 1.39 (0.83, 2.34) |
| 50-54 vs 40-45 | 2.51 (1.58, 3.98) | 1.85 (1.73, 1.97) | 2.11 (1.32, 3.39) |
| 55-59 vs 40-45 | 3.72 (2.40, 5.77) | 2.64 (2.48, 2.81) | 3.54 (2.27, 5.52) |
| 60-64 vs 40-45 | 6.03 (3.94, 9.22) | 3.43 (3.23, 3.65) | 6.32 (4.11, 9.72) |
| 65-69 vs 40-45 | 8.63 (5.66, 13.14) | 4.37 (4.11, 4.64) | 12.54 (8.20, 19.19) |
| 70-74 vs 40-45 | 14.38 (9.49, 21.80) | 5.53 (5.21, 5.87) | 22.24 (14.59, 33.90) |
| 75-79 vs 40-45 | 19.99 (13.19, 30.30) | 7.73 (7.28, 8.20) | 33.32 (21.85, 50.81) |
| 80-84 vs 40-45 | 31.45 (20.78, 47.61) | 11.09 (10.45, 11.77) | 53.74 (35.27, 81.86) |
| 85+ vs 40-45 | 65.32 (43.28, 98.59) | 22.85 (21.55, 24.22) | 83.68 (55.14, 126.99) |
| Male vs female | 1.72 (1.61, 1.84) | 1.39 (1.37, 1.41) | 1.88 (1.74, 2.03) |
| Reside in rural vs urban^e^ | 0.72 (0.59, 0.87) | 1.02 (0.99, 1.04) | 0.91 (0.72, 1.14) |
| Asthma | 1.09 (1.00, 1.19) | 0.90 (0.88, 0.92) | 1.00 (0.91, 1.11) |
| Chronic obstructive pulmonary disease | 1.34 (1.22, 1.48) | 1.63 (1.60, 1.66) | 1.34 (1.19, 1.50) |
| Hypertension | 1.36 (1.24, 1.50) | 1.07 (1.06, 1.09) | 1.29 (1.16, 1.44) |
| Diabetes | 1.41 (1.32, 1.51) | 1.13 (1.12, 1.15) | 1.17 (1.08, 1.27) |
| Congestive heart failure | 1.29 (1.19, 1.41) | 1.60 (1.57, 1.63) | 1.13 (1.02, 1.25) |
| Dementia or frailty score >15 | 3.23 (2.99, 3.50) | 2.15 (2.11, 2.19) | 1.66 (1.51, 1.82) |
| Cancer^f^ | 1.29 (1.13, 1.48) | 3.60 (3.53, 3.67) | 1.94 (1.65, 2.27) |
| Chronic kidney disease^f^ |  |  |  |
| With no recent dialysis vs no disease | 1.54 (1.42, 1.67) | 1.24 (1.21, 1.26) | 1.83 (1.66, 2.01) |
| With recent dialysis vs no disease | 3.54 (2.86, 4.39) | 2.72 (2.59, 2.86) | 2.76 (2.11, 3.61) |
| Immunocompromised^g^ | 1.50 (1.26, 1.79) | 1.67 (1.62, 1.73) | 1.35 (1.07, 1.70) |
| Advanced Liver Disease | 1.17 (0.96, 1.42) | 2.16 (2.10, 2.23) | 1.55 (1.25, 1.92) |
| Cardiac ischemic disease | 0.91 (0.83, 0.99) | 0.90 (0.88, 0.92) | 0.96 (0.86, 1.06) |
| Ischemic stroke or transient ischemic attack^h^ | 1.23 (1.10, 1.38) | 1.16 (1.13, 1.19) | 1.02 (0.89, 1.16) |
| Hospital admission, past 3 years |  |  |  |
| Once vs 0 times | 1.63 (1.49, 1.77) | 2.03 (2.00, 2.07) | 1.47 (1.33, 1.63) |
| Twice vs 0 times | 2.22 (1.99, 2.48) | 2.69 (2.63, 2.76) | 1.71 (1.50, 1.95) |
| Three times or more vs 0 times | 2.32 (2.06, 2.61) | 3.45 (3.37, 3.53) | 1.89 (1.65, 2.16) |
| Outpatient physician visits, past year |  |  | 1.47 (1.33, 1.63) |
| 2-4 times vs 0-1 times | 1.47 (1.28, 1.69) | 1.02 (0.99, 1.04) | 0.90 (0.79, 1.02) |
| 5-8 times vs 0-1 times | 1.50 (1.32, 1.72) | 1.03 (1.00, 1.05) | 1.03 (0.90, 1.16) |
| 9-14 times vs 0-1 times | 1.47 (1.29, 1.68) | 1.11 (1.08, 1.14) | 1.07 (0.93, 1.22) |
| 15 times or more vs 0-1 times | 1.70 (1.48, 1.95) | 1.54 (1.50, 1.58) | 1.09 (0.94, 1.26) |
| Income (1= Highest)^i,j^ |  |  |  |
| 2 vs 1 | 1.13 (1.00, 1.29) | 1.01 (0.98, 1.03) | 1.07 (0.92, 1.24) |
| 3 vs 1 | 1.32 (1.16, 1.51) | 1.05 (1.02, 1.07) | 1.20 (1.03, 1.40) |
| 4 vs 1 | 1.28 (1.10, 1.48) | 1.07 (1.04, 1.10) | 1.25 (1.06, 1.49) |
| 5 vs 1 | 1.30 (1.09, 1.54) | 1.16 (1.12, 1.20) | 1.16 (0.94, 1.42) |
| Educational attainment (1=Lowest)^i,k^ |  |  |  |
| 2 vs 1 | 1.16 (1.04, 1.30) | 1.04 (1.02, 1.07) | 0.96 (0.85, 1.10) |
| 3 vs 1 | 1.10 (0.97, 1.24) | 1.06 (1.03, 1.09) | 1.01 (0.88, 1.17) |
| 4 vs 1 | 1.21 (1.06, 1.38) | 1.09 (1.06, 1.12) | 1.02 (0.88, 1.19) |
| 5 vs 1 | 1.27 (1.10, 1.47) | 1.14 (1.11, 1.18) | 1.06 (0.89, 1.25) |
| Proportion essential workers (1=Lowest) ^i,l^ |  |  |  |
| 2 vs 1 | 1.12 (1.00, 1.26) | 1.04 (1.02, 1.07) | 1.01 (0.88, 1.16) |
| 3 vs 1 | 1.17 (1.03, 1.33) | 1.08 (1.05, 1.11) | 1.05 (0.91, 1.23) |
| 4 vs 1 | 1.26 (1.10, 1.45) | 1.06 (1.03, 1.09) | 1.10 (0.93, 1.30) |
| 5 vs 1 | 1.28 (1.10, 1.50) | 1.12 (1.09, 1.16) | 1.09 (0.91, 1.31) |
| Proportion racialized groups (1=Lowest) ^i,m^ |  |  |  |
| 2 vs 1 | 1.25 (1.06, 1.46) | 0.98 (0.96, 1.00) | 1.08 (0.90, 1.31) |
| 3 vs 1 | 1.37 (1.16, 1.61) | 0.99 (0.97, 1.02) | 1.02 (0.84, 1.23) |
| 4 vs 1 | 1.46 (1.22, 1.74) | 0.94 (0.92, 0.97) | 1.04 (0.84, 1.27) |
| 5 vs 1 | 1.42 (1.16, 1.73) | 0.88 (0.85, 0.92) | 1.08 (0.85, 1.36) |
| Proportion recent immigrants (1=Lowest) ^i,n^ |  |  |  |
| 2 vs 1 | 1.07 (0.97, 1.17) | 0.98 (0.96, 1.00) | 1.05 (0.94, 1.18) |
| 3 vs 1 | 1.06 (0.95, 1.17) | 0.96 (0.94, 0.99) | 1.08 (0.95, 1.22) |
| Proportion apartment buildings (1=Lowest) ^i,o^ |  |  |  |
| 2 vs 1 | 1.13 (1.02, 1.25) | 1.02 (1.00, 1.04) | 0.99 (0.88, 1.11) |
| 3 vs 1 | 1.25 (1.11, 1.41) | 1.00 (0.97, 1.03) | 1.04 (0.90, 1.20) |
| Average household size (1=Lowest) ^i,p^ |  |  |  |
| 1 vs 3 | 1.11 (0.98, 1.27) | 1.06 (1.04, 1.09) | 1.07 (0.92, 1.25) |
| 2 vs 3 | 0.97 (0.85, 1.10) | 1.05 (1.02, 1.07) | 1.08 (0.93, 1.26) |
| 4 vs 3 | 1.16 (1.03, 1.31) | 0.97 (0.95, 1.00) | 1.04 (0.90, 1.20) |
| 5 vs 3 | 1.30 (1.13, 1.48) | 0.85 (0.83, 0.88) | 1.01 (0.86, 1.19) |
| Proportion high-density housing (1=Lowest) ^i,q^ |  |  |  |
| 2 vs 1 | 0.92 (0.84, 1.02) | 1.02 (1.00, 1.04) | 0.90 (0.80, 1.02) |
| 3 vs 1 | 0.90 (0.81, 1.00) | 1.04 (1.02, 1.07) | 0.91 (0.81, 1.03) |
| 4 vs 1 | 1.01 (0.90, 1.13) | 1.07 (1.04, 1.09) | 0.82 (0.72, 0.94) |
| Public health region |  |  |  |
| CentralEast vs Toronto | 0.35 (0.28, 0.44) | 1.09 (1.06, 1.13) | 1.03 (0.80, 1.35) |
| CentralWest vs Toronto | 0.68 (0.61, 0.77) | 1.12 (1.09, 1.15) | 0.82 (0.72, 0.95) |
| Durham vs Toronto | 0.51 (0.42, 0.63) | 1.16 (1.12, 1.21) | 0.77 (0.61, 0.97) |
| Eastern vs Toronto | 0.17 (0.13, 0.24) | 1.12 (1.08, 1.16) | 0.59 (0.41, 0.84) |
| North vs Toronto | 0.19 (0.14, 0.25) | 1.19 (1.14, 1.23) | 0.84 (0.57, 1.24) |
| Ottawa vs Toronto | 0.53 (0.45, 0.63) | 1.12 (1.09, 1.16) | 0.98 (0.80, 1.20) |
| Peel vs Toronto | 0.91 (0.81, 1.01) | 1.02 (0.98, 1.05) | 0.87 (0.76, 0.99) |
| SouthWest vs Toronto | 0.73 (0.64, 0.84) | 1.11 (1.07, 1.14) | 0.94 (0.80, 1.11) |
| York vs Toronto | 0.83 (0.73, 0.94) | 0.94 (0.91, 0.98) | 0.87 (0.75, 1.01) |
| Year and month of COVID-19 diagnosis |  |  |  |
| 2020-03 vs 2020-08 |  |  | 6.29 (3.45, 11.47) |
| 2020-04 vs 2020-08 |  |  | 4.28 (2.39, 7.67) |
| 2020-05 vs 2020-08 |  |  | 2.76 (1.52, 5.01) |
| 2020-06 vs 2020-08 |  |  | 1.66 (0.86, 3.21) |
| 2020-07 vs 2020-08 |  |  | 1.54 (0.76, 3.14) |
| 2020-09 vs 2020-08 |  |  | 1.46 (0.77, 2.75) |
| 2020-10 vs 2020-08 |  |  | 1.56 (0.86, 2.83) |
| 2020-11 vs 2020-08 |  |  | 1.67 (0.93, 3.00) |
| 2020-12 vs 2020-08 |  |  | 2.23 (1.25, 3.97) |
| 2021-01 vs 2020-08 |  |  | 2.27 (1.28, 4.05) |

^a^Death within 30 days following or 7 days prior to a lab-confirmed positive SARS-CoV-2 test was considered COVID-19-related;

^b^Death without a lab-confirmed positive SARS-CoV-2 test;

^c^Death within 30 days following or 7 days prior to a lab-confirmed positive SARS-CoV-2 test among those diagnosed with COVID-19;

^d^Cause-specific hazard models were used for COVID-19-related mortality analyses, non-COVID-19 mortality analyses; and logistic regression models were used for COVID-19 case fatality analyses;

^e^We defined rural as being located outside the commuting zone of a city with a population greater than 10 000;

^f^Diagnosis in the last 5 years;

^g^Immunocompromised defined as diagnosed with HIV, had an organ or bone marrow transplant, or had another immunodeficient condition;

^h^Diagnosis in the last 20 years;

^i^Area-level variables at the level of the Census Dissemination Area (DA), which usually comprises 400-700 individuals;

^j^Income quintile has variable cut-of values in each city or Census area, to take cost of living into account; A DA being in quintile 1 means it is among the highset 20% of dissenmination areas in its city by median household income;

^k^1^st^ quintile represents DAs with 17.1%–94.3% of people aged 25–64 years without a diploma; 2^nd^ quintile, 11.4%–17.1% of people; 3^rd^ quintle, 7.5%–11.4% of people; 4^th^ quintile, 4.1%–7.5% of people; and 5^th^ quintile, 0%–4.1% of people;

^l^1^st^ quintile represents 0%–32.5% of working people in the area who self-identified as working in an essential job, including sales, trades, manufacturing, and agriculture; 2^nd^ quintile, 32.5%–42.3% of people; 3^rd^ quintile, 42.3%–49.8% of people; 4^th^ quintile, 50.0%–57.5% of people; and 5^th^ quintile, 57.5%–114.3% of people;

^m^1^st^ quintile represents 0%–2.2% of people in the area who self-identified as a visible minority; 2^nd^ quintile, 2.2%–7.5% of people; 3^rd^ quintile: 7.5%–18.7% of people; 4^th^ quintile, 18.7%–43.5% of people; and 5^th^ quintile, 43.5%–100% of people;

^n^1^st^ category represents 0%–2.1% of people in the area being recent immigrants who came to Canada within the last 5 years; 2^nd^ category, 2.1%–4.7% of people; and 3^rd^ category, 4.7%–41.2% of people;

^o^1^st^ category, 0%–7.3% of buildings in the area are apartment buildings; 2^nd^ category, 7.4%–37.7% are apartment buildings; and 3^rd^ category, 37.7%–100% are apartment buildings;

^p^1^st^ quintile represents 0–2.1 people/dwelling; 2^nd^ quintile, 2.2–2.4 people/dwelling; 3^rd^ quintile, 2.5–2.6 people/dwelling; 4^th^ quintile, 2.7–3 people/dwelling; and 5^th^ quintile, 3.1–5.7 people/dwelling;

^q^1^st^ category represents 0–2.6% of households are considered high-density housing; 2^nd^ category, 2.7-5.2%; 3^rd^ category, 5.3-8.7%; 4^th^ category, >8.7%; the high frequency of zeros permitted the creation of only 4 categories (the lower 2 quintiles combined);Abbreviation: IQR: interquartile range;

Abbreviations: CI, confidence interval.

**Appendix Table 5.** Social determinants of health (SDOH) in COVID-19-related mortality^a^, stratified by residence in rural vs. urban area among community dwelling adult populations aged 20 years and older in Ontario, Canada, March 1 2020 – Mar 2, 2021.

|  | Fully adjusted hazard ratio (95% confidence interval) | |
| --- | --- | --- |
| Area-level SDOH^b^ | **Urban** | **Rural** |
| Income quintile (1= Highest) |  |  |
| 2 vs 1 | 1.18 (1.04, 1.35) | 0.87 (0.50, 1.52) |
| 3 vs 1 | 1.39 (1.22, 1.59) | 0.73 (0.40, 1.34) |
| 4 vs 1 | 1.36 (1.18, 1.57) | 0.94 (0.49, 1.80) |
| 5 vs 1 | 1.42 (1.20, 1.68) | 0.56 (0.25, 1.27) |
| Educational attainment quintile (1=Lowest) |  |  |
| 2 vs 1 | 1.20 (1.07, 1.34) | 0.82 (0.35, 1.89) |
| 3 vs 1 | 1.14 (1.01, 1.29) | 0.74 (0.33, 1.68) |
| 4 vs 1 | 1.25 (1.10, 1.42) | 0.86 (0.38, 1.95) |
| 5 vs 1 | 1.31 (1.14, 1.51) | 1.09 (0.47, 2.54) |
| Proportion essential workers quintile (1=Lowest) |  |  |
| 3 vs 1,2 combined | 1.06 (0.96, 1.17) | 1.62 (0.87, 3.02) |
| 4 vs 1,2 combined | 1.16 (1.04, 1.30) | 1.18 (0.61, 2.28) |
| 5 vs 1,2 combined | 1.13 (0.99, 1.29) | 2.01 (1.04, 3.90) |
| Proportion racialized groups quintile (1=Lowest) |  |  |
| 3 vs 1,2 combined | 1.27 (1.06, 1.52) | 0.95 (0.65, 1.40) |
| 4,5 combined vs 1,2 combined | 1.41 (1.19, 1.68) | 1.46 (0.71, 2.99) |
| Proportion recent immigrants (1=Lowest) |  |  |
| 2,3 combined vs 1 | 1.07 (0.98, 1.16) | 1.05 (0.56, 1.98) |
| Proportion apartment buildings (1=Lowest) |  |  |
| 2 vs 1 | 1.12 (1.01, 1.24) | 1.63 (1.04, 2.57) |
| 3 vs 1 | 1.20 (1.06, 1.36) | 5.68 (2.36, 13.64) |
| Average household size (1=Lowest) |  |  |
| 1 vs 3 | 1.17 (1.03, 1.34) | 0.28 (0.14, 0.56) |
| 2 vs 3 | 1.03 (0.90, 1.18) | 0.40 (0.25, 0.66) |
| 4 vs 3 | 1.22 (1.08, 1.38) | 0.71 (0.43, 1.16) |
| 5 vs 3 | 1.36 (1.19, 1.55) | 0.77 (0.35, 1.72) |
| Proportion high-density housing (1=Lowest) |  |  |
| 2 vs 1 | 0.92 (0.83, 1.02) | 1.02 (0.67, 1.56) |
| 3 vs 1 | 0.88 (0.80, 0.98) | 1.16 (0.70, 1.92) |
| 4 vs 1 | 1.00 (0.89, 1.11) | 1.74 (0.85, 3.55) |

^a^Death within 30 days following or 7 days prior to a lab-confirmed positive SARS-CoV-2 test was considered COVID-19-related;

^b^All area-level SDOH variables are measured at the level of the Census Dissemination Area, and detailed definitions of these variables are shown in **Table 1** footnotes.
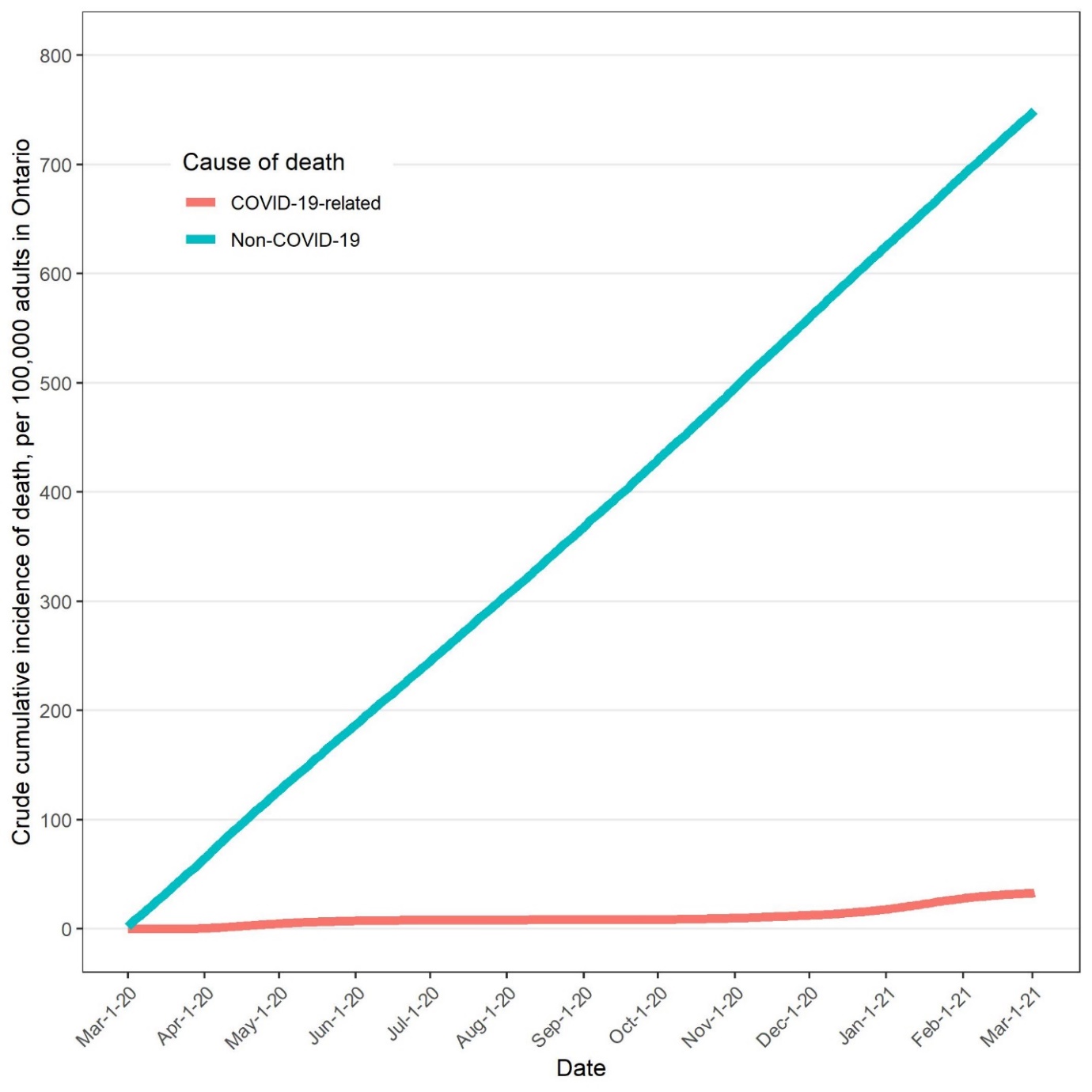

**Appendix Figure 1. Cumulative incidence functions for COVID-19-related mortality and non-COVID-19 mortality among community dwelling adult populations aged 20 years and older in Ontario, Canada, March 1 2020 – Mar 2, 2021.** COVID-19-related mortality defined as death within 30 days following or 7 days prior to a positive SARS-CoV-2 test; non-COVID-19 mortality defined as death without a positive SARS-CoV-2 test.

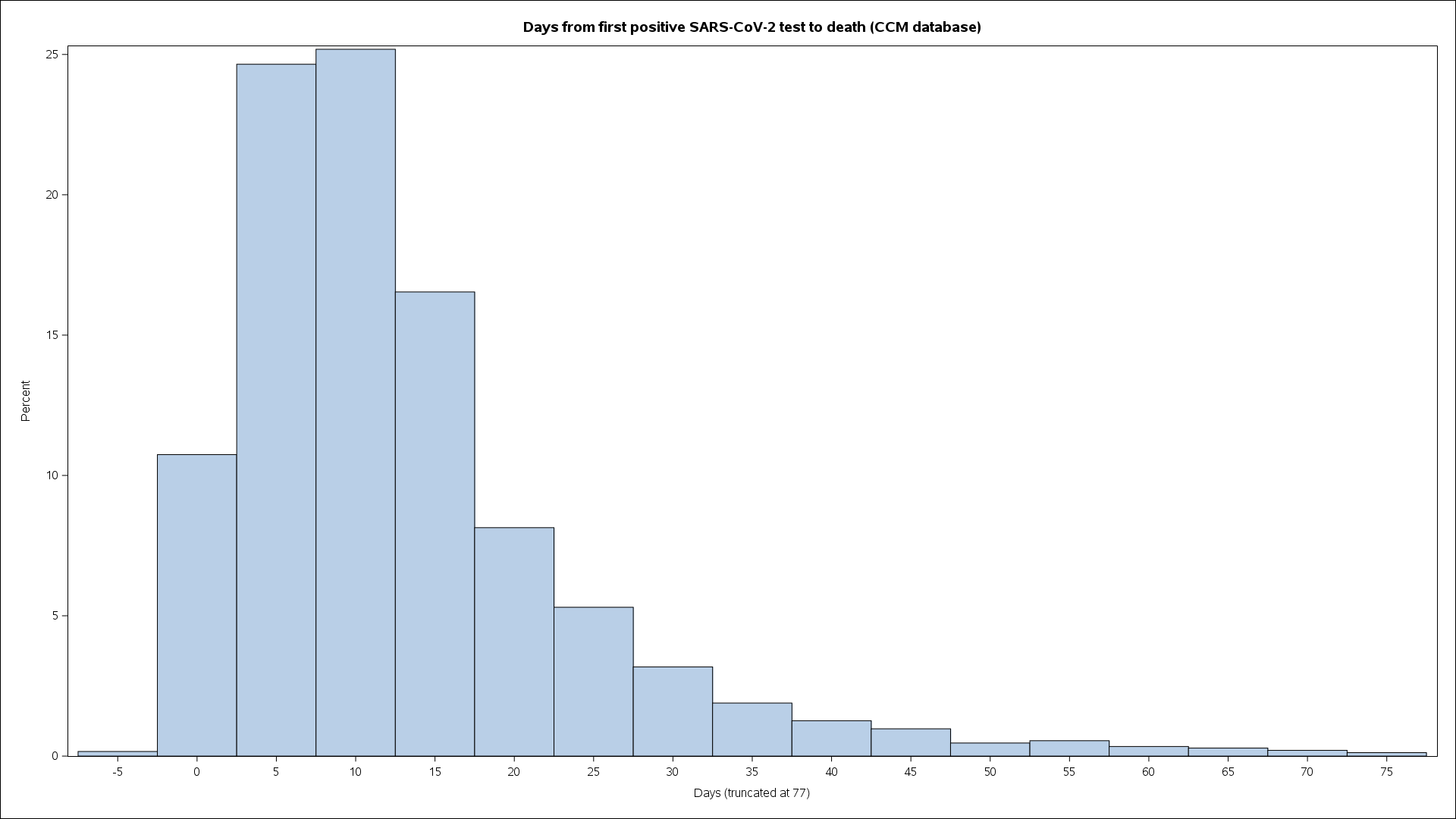

**Appendix Figure 2. Distribution of time from COVID-19 diagnosis to all-cause death.** Of all deaths occurred and recorded in the CCM databases among individuals diagnosed with COVID-19, 92% occurred within 30 days following or 7 days prior to a positive test. CCM: COVID-19 Case and Contact Management System.
